# Increased aperiodic neural activity during sleep in major depressive disorder

**DOI:** 10.1101/2022.06.03.22275735

**Authors:** Yevgenia Rosenblum, Leonore Bovy, Frederik D. Weber, Axel Steiger, Marcel Zeising, Martin Dresler

## Abstract

**Background:** In major depressive disorder (MDD), patients often express subjective sleep complaints while polysomnographic studies report only subtle alterations in neural oscillations. We hypothesize that the study of aperiodic electroencephalographic (EEG) dynamics, a marker of excitation-to-inhibition balance, may bring new insights into our understanding of sleep abnormalities in MDD. Specifically, we investigate aperiodic neural activity during sleep and its relationships with the time of sleep, depression severity, and responsivity to antidepressant treatment.

**Methods:** Polysomnography was recorded in 38 MDD patients (in unmedicated and 7d medicated states) and 38 age-matched healthy controls (N1=76). Aperiodic EEG activity was evaluated using the Irregularly Resampled Auto-Spectral Analysis with slopes’ means and intra-individual variability as outcome measures. Depression severity was assessed with the Hamilton Depression Rating Scale. We replicated the analysis using two independently collected datasets of medicated patients and controls (N2=60, N3=80).

**Results:** Unmedicated patients showed flatter aperiodic slopes compared to controls during N2 (p-value=0.002) and steeper slopes compared to their later medicated state (p-values<0.02) during all sleep stages. Within unmedicated patients, slopes were flatter during late compared to early N2 sleep (p-value=0.006). Late N2 slopes further correlated with depression severity after 7d of antidepressant treatment (r=-0.34, p-value=0.04). Variability of slopes was increased in both unmedicated (p-values<0.03) and medicated states (p-values <0.006) of patients’ N2, N3, and REM sleep compared to controls.

**Conclusion:** Flatter slopes of aperiodic EEG power with increased variability may reflect unstable, noisy neural activity due to increased excitation-to-inhibition balance, representing a new disease-relevant feature of sleep in MDD.

## 1. Introduction

Major depressive disorder (MDD) is a common psychiatric disorder characterized by at least two weeks of pervasive low mood, anhedonia, inappropriate guilt, and feelings of worthlessness (1). In 2017, MDD affected ∼2% of the world population (2). The percentage of people who are affected at one point in their life varies from 7% to 21%, reflecting the fact that MDD is a serious public health problem (2). Besides abnormalities of mood and affect, MDD patients often have sleep complaints, including insomnia (in ∼60%) or hypersomnia (in ∼15%), as well as fatigue, excessive daytime sleepiness, and lack of concentration while awake (3). Broad evidence suggests that disturbances of sleep-wake rhythms and circadian time-keeping system underlie the pathophysiology of depression. Understanding the mechanisms of these alterations might bring new insights into the understanding of MDD.

Intriguingly, whereas some polysomnographic studies confirm subjective sleep complaints of the patients by reporting decreased slow-wave and delta amplitudes, higher spindle amplitude, lower spindle density, and a more dispersed slow-wave-spindle coupling, others suggest that electroencephalographic (EEG) changes in MDD might be more subtle (4, 5). Thus far, however, EEG analysis in MDD has been limited to neural oscillations whereas aperiodic (i.e., non-oscillatory, 1/f) dynamics have not been explored.

Aperiodic activity is a distinct and functionally significant type of brain dynamics, which currently receives increasing attention (6, 7). Aperiodic dynamics display an asynchronous input and follow a power-law function with a 1/f-like shape, where power decreases with increasing frequency (8). Historically, aperiodic activity was often interpreted as noise or simple summation of many oscillations and, therefore, underexplored (9-11). Recently, however, it has been shown that the slope of the aperiodic component reflects the ratio between excitatory and inhibitory currents in the brain (12). Given the crucial role of the proper balance between neural excitation and inhibition (E/I) for healthy cognition, behavior (13) and sleep, aperiodic activity seems to be a promising tool for investigating MDD with its cholinergic, monoaminergic (14, 15), glutamatergic (16) and GABAergic imbalance (17, 18). In MDD, the E/I ratio could be further affected by prescribed antidepressants.

Based on this background, we hypothesize that aperiodic dynamics could bring new insights into the pathophysiology of sleep in MDD and explore aperiodic activity during sleep as well as its relationships with the time of sleep, depression severity, and responsivity to antidepressant treatment. We expect to observe some changes in aperiodic slopes (reflecting altered E/I balance), however, without a prior hypothesis on the direction of such changes, as well as increased intra-individual variability of slopes (indicating decreased state stability due to altered E/I balance) in patients compared to controls.

## 2. Methods and Materials

### Participants

We retrospectively analyzed polysomnographic recordings from a previous study conducted at the Max Planck Institute of Psychiatry, Munich, Germany (5). The sample consisted of 40 patients with MDD and 40 healthy controls individually matched by age (±2 years of tolerance) and gender (See reference 5 for details). Exclusion criteria included suicidality, shift working, transmeridian flights in the preceding three months, drug or alcohol dependence, professional piano skills, professional typewriting skills, sleep disorders, pregnancy, and a history of severe physical disorders. Subjects who received long-acting medication before the beginning of the experiment were excluded if the treatment was not stopped in time to ensure a complete wash-out (e.g., antipsychotics, fluoxetine). Due to technical failure in the EEG data of two medicated patients, all paired analyses were matched on the remaining full datasets (n=38 per group). To confirm the results, we replicated the analyses using two independently collected datasets of short and long-term medicated MDD patients and age and gender-matched healthy controls (Supplementary Material S4). All studies were approved by the Ethics committee of the University of Munich. All participants gave written informed consent.

### Questionnaires

Depression severity of patients was measured with the Hamilton Depression Rating Scale (HAM-D) at baseline (“unmedicated”) and 7 days after the commencement of antidepressant treatment (“medicated”). A higher score reflects higher depression severity. In Supplementary Material S3, we also report the Pittsburgh Sleep Quality Index, which was available in a subset of the patients.

### Polysomnography

All participants slept in the sleep laboratory, and all had an adaptation night before the examination night. For the EEG of the examination night, 118 Ag/AgCl electrodes were applied using an Easycap 128Ch-BrainCap (EasyCap GmbH, Herrsching, Germany). Polysomnography was recorded (sampling rate of 200 Hz), stored, and digitized following the 10-5 system with a JE-209A amplifier (Neurofax Software, Nihon Kohden Europe GmbH, Rosbach, Germany) with a common-mode rejection ratio of ≥ 110 dB and with impedances below 10 kOhm, including EEG (filtered at 0.016 Hz high pass only, -6 dB/octave), electrooculography, mental/submental electromyography with a ground electrode attached at the forehead. For the EEG, the average of the electrodes AFF5H and AFF1H was defined as the reference.

Polysomnography of the patients was recorded at two timepoints: when unmedicated and when 7d medicated. Sleep was scored by independent experts according to the AASM standards (19). All epochs scored as wake before the sleep onset and after morning awakening were rejected. Epochs with artifacts were rejected based on visual inspection. In Supplementary Material S2, we also report morning resting-state EEG measured in a subset of participants to explore whether the observed effects are specific to sleep.

### Aperiodic activity

The aperiodic (fractal) component of the EEG power was calculated with the Irregularly Resampled Auto-Spectral Analysis (20) for each 30 s of sleep, corresponding to the conventionally defined sleep epochs. A MATLAB implementation of the algorithm was adapted from the Fieldtrip tutorial website (http://www.fieldtriptoolbox.org/example/irasa/), using the *ft_freqanalysis* function with *cfg*.*taper=‘hanning’* and *cfg*.*method=‘irasa’*. Aperiodic power was transformed to log-log coordinates by standard least squares regression, where the slope of the line was calculated as the power-law exponent estimation.

The signal was filtered in the 0.2–48 Hz frequency band. In Supplementary Material S1, we also report low (2–20 Hz) and high (30–48 Hz) bands analyses to control for a possible distortion of the linear fit by excluding low frequencies with strong oscillatory activity (12) and for the reliable discrimination between wakefulness and REM sleep, respectively (21).

The slopes were averaged over each sleep stage as defined by the hypnogram over five topographical areas: 1) frontal (Fz, F1, F2, F3, F4, F5, F6, F7, F8, F9, F10); 2) central (Cz, C1, C2, C3, C4, C5, C6); 3) parietal (Pz, P1, P2, P3, P4, P5, P6, P7, P8, P9, P10); 4) occipital (Oz, O1, O2); 5) temporal (T7, T8). To assess the degree of neural noise we also calculated intra-individual variability of slopes as a standard deviation over all epochs of a given stage and area in each participant and then averaged these values over all participants of a given group.

### Time

Besides the effect of a sleep stage on aperiodic slopes, we also studied the simultaneous effect of the sleep stage and time within the sleep period on aperiodic slopes. This exploratory sub-analysis assessed whether the effect of sleep stages is entirely related to the influence of sleep or can be partly attributable to the circadian system. To answer this question, we averaged the slopes of the aperiodic power component over the early (0.5–2 h of the sleep period) and late (4.5–6 h of the sleep period) non-REM 2 epochs separately. 6 h was chosen as the latest time point for which the data was available for all participants, i.e., some participants did not sleep more than 6 h. Unfortunately, we could not stratify other sleep stages in this way as we had not enough (only 0– 7 epochs) evening or morning epochs for the comparison.

### Medication

To assess specific medication effects, we stratified the patients by antidepressant class, namely, tricyclic antidepressants (TCAs, n=8), selective serotonin reuptake inhibitors (SSRIs, n=13), norepinephrine–dopamine reuptake inhibitor (NDRI, n=6), or serotonin-norepinephrine reuptake inhibitors (SNRI, n=6). Only 5 patients took noradrenergic and specific serotonergic antidepressants (NaSSA); therefore, this group was not analyzed separately. None of the patients was treated with sedative antidepressants. In addition, we stratified the patients by the REM-suppressive (n=21) vs REM-non-suppressive (n=17) antidepressants.

### Statistical analysis

Firstly, aperiodic activity was analyzed using three-way ANCOVA with the five-level “brain area” and five-level “sleep stage” as within-subject factors. The “study group” served as a two-level between-subjects factor to compare 1) unmedicated patients and controls; 2) the same patients when 7d medicated and controls. Even though we matched the participants’ age individually, given that at the group level the age ranged from 19 to 54 years, we added to the analysis the “age” factor as a covariate. We performed an additional ANOVA to compare 3) unmedicated and 7d medicated states of the patients using the “brain area”, “sleep stage”, and “state” as within-subject factors.

Secondly, we performed five two-way ANCOVAs for each sleep stage separately with the “brain area”, “study group” and “age” factors to compare patients and controls and five two-way ANOVAs with the “brain area” and “state” factors to compare unmedicated and medicated states of the patients. The Benjamini-Hochberg’s adjustment was applied to control for multiple comparisons with a false discovery rate set at 0.05. Due to the semi-exploratory nature of this study, all corrections were done per research question (5 tests reflecting the number of sleep stages) with the α-level set in the 0.01–0.05 range. For all ANOVAs/ANCOVAs we applied Greenhouse-Geisser correction since Mauchly’s test revealed that the sphericity assumption was violated (ε<0.75, p<0.05). The assumptions of normality and homogeneity of variance were tested using the Q-Q plot and Levene’s homogeneity test, respectively.

Thirdly, we performed *post hoc* analysis to compare each pair of groups for each area and sleep stage separately. We used the two-tailed Student’s unpaired t-test to compare patients to controls and paired t-test to compare the unmedicated and medicated states of the patients. Effect sizes were calculated with Cohen’s d.

To study the effect of time, we performed two-way ANOVAs with the five-level “brain area” and two-level “time” (early vs late sleep) within-subject factors in each group separately. Then, we performed *post hoc* analysis using the paired t-test and Cohen’s d to compare early and late sleep for each area of interest in each group separately.

To study the effect of the antidepressant treatment we used the non-parametric Mann-Whitney U two-tailed test as after stratifying the patients by antidepressant classes the samples became rather small. Benjamini-Hochberg’s adjustment for 25 tests (5 stages by 5 areas) was applied with the α-level set in the 0.002–0.050 range.

The diagnostic accuracy of the aperiodic slopes was defined using the area under the Receiver Operating Characteristic curve (AUC). Correlations between means of the frontal aperiodic slopes and HAM-D scores of the participants at baseline and 7d were assessed with Pearson’s correlation coefficients. SPSS software (version 25; SPSS, Inc) was used for all statistical analyses.

## 3. Results

The demographic, clinical, and sleep characteristics of the participants are reported in our previous paper, where this dataset is referred to as “Dataset B” (5).

### 3.1. Means of slopes

#### Unmedicated patients vs controls

The three-way ANCOVA revealed a main effect of the sleep stage (F=82.2, p<0.001) on the slopes without interactions. The two-way ANCOVA revealed a main effect of the group (without a group-area interaction) on the slopes measured during the N2 stage (the obtained p=0.002 < the corrected threshold p=0.010). The main effect during N3 sleep did not pass the correction for multiple comparisons (the obtained p=0.037 > the corrected threshold p=0.020).

The *post hoc* analysis revealed that unmedicated patients showed flatter slopes compared to the controls during N2 sleep in all areas and during N3 sleep – in the frontal and temporal areas with moderate to large effect sizes. Slopes of the wake, N1, and REM epochs were comparable in both groups (Fig.1, Table 1).

**Figure 1.**
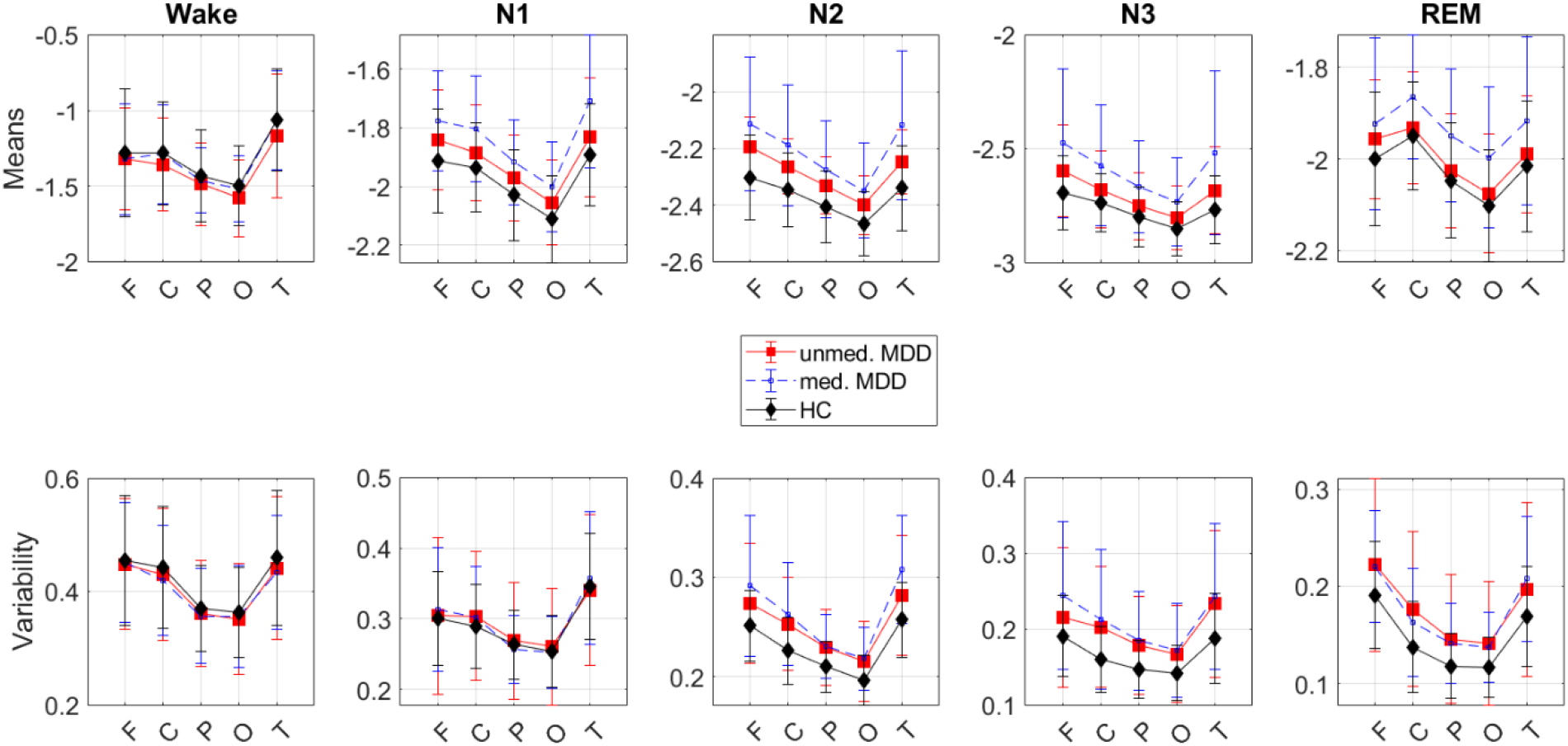
Means and variability of aperiodic slopes. The means **(top)** and intra-individual variability **(bottom)** of the slopes of the broadband (0.2–48 Hz) aperiodic power component over each sleep stage, area of interest and study group. **Top:** Unmedicated patients (red) show flatter (more positive values) slopes during N2 compared to controls (black) in all areas. The flattest slopes are observed in the frontal and temporal areas. 7d medicated patients (blue) show flatter slopes compared to their own unmedicated state (red) and controls (black) during all sleep stages – but not wake epochs – in all areas. **Bottom:** Patients in both unmedicated and 7d medicated states show greater intra-individual slopes variability during N2, N3, and REM compared to controls. MDD – 38 major depressive disorder patients, unmed. – unmedicated, med. – 7d medicated MDD patients, HC – 38 healthy controls, F – frontal, C – central, P – parietal, O – occipital, T – temporal electrodes.

**Table 1:**
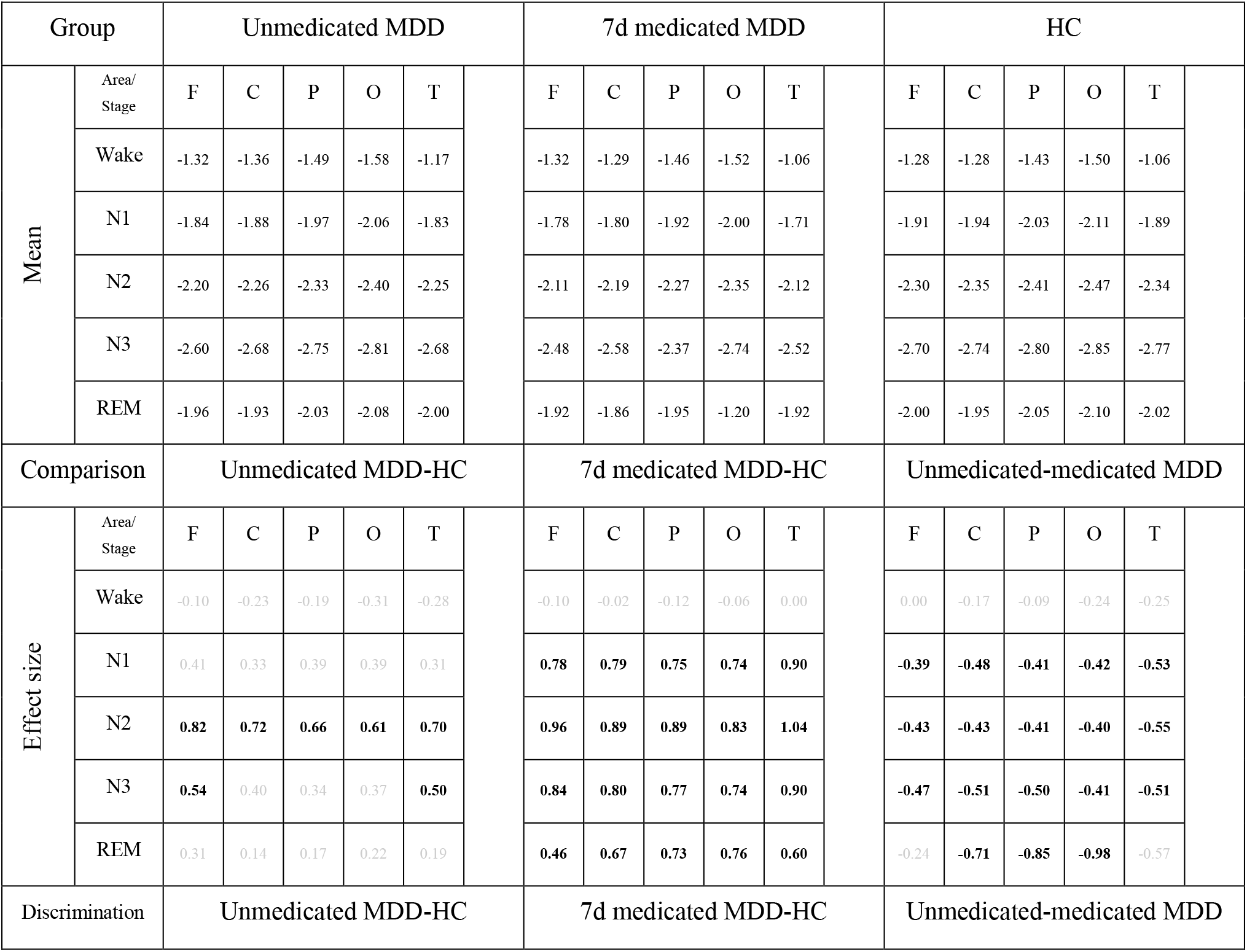
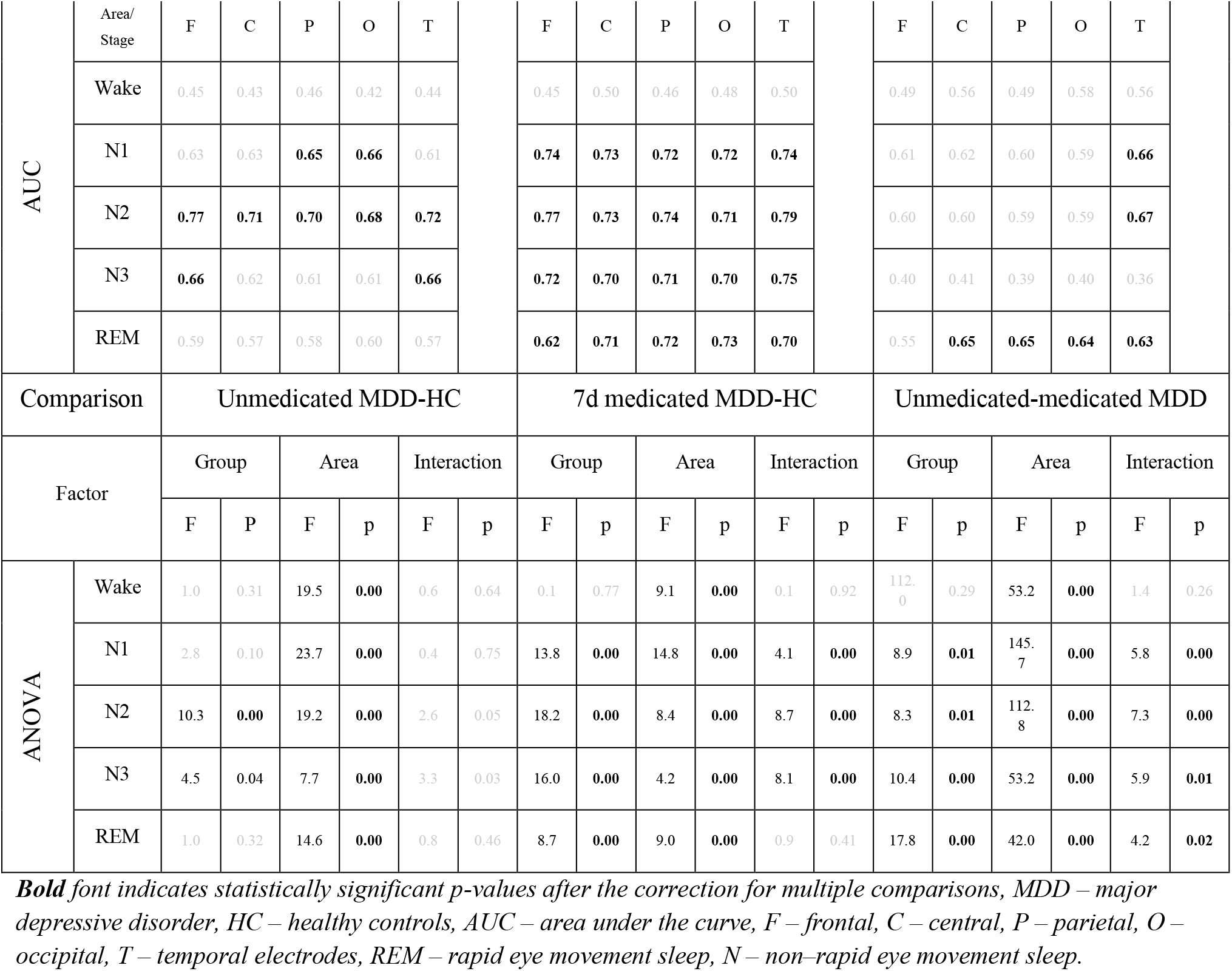
Slope means.

#### Medicated patients vs controls

The three-way ANCOVA revealed area-group (F=2.8, p=0.04), stage-group (F=4.7, p=0.01), and area-stage (F=6.6, p<0.001) interactions. The two-way ANCOVA revealed an effect of the group on the slopes measured during N1, N2, and N3 sleep (all p-values<0.001) with a significant area-group interaction and during REM sleep (p=0.004) – without an interaction.

The *post hoc* analysis revealed that the difference between the groups was detectable in all areas with moderate to large effect sizes. During non-REM sleep, the difference between the groups was greater in the frontal and temporal electrodes compared to other areas. Slopes of the wake epochs were comparable (Fig.1, Table 1).

These findings were replicated using two independently collected datasets of short and long-term medicated patients (Supplementary Material S4).

#### Unmedicated vs medicated states

The three-way ANOVA revealed area-state (F=4.0, p=0.012) and area-stage (F=27.5, p<0.001) interactions. The two-way ANCOVA revealed a main effect of the N1, N2, N3, and REM stages (all p-values<0.007) with an area-state interaction.

The *post hoc* analysis revealed that patients showed flatter slopes when medicated than when unmedicated during N1, N2, and N3 sleep in all areas (all p-values<0.02) with moderate effect sizes and during REM sleep – in the central, parietal, and occipital areas (all p-values<0.001) with moderate-to-large effect sizes. During the N1 and N2 stages, the difference between the states was greater in the temporal electrodes compared to other areas, and during REM sleep – in the occipital electrodes compared to other areas and in the parietal electrodes compared to the central, frontal, and temporal areas. Slopes of the wake epochs were comparable (Fig.1, Table 1).

#### Within-group topographical effect

There was a significant within-subject topographical effect on the slopes during all sleep stages. Specifically, during the N1, N2, and N3 stages, the participants showed flatter slopes in the frontal and temporal electrodes compared to other areas, whereas during REM sleep, they showed flatter slopes in the frontal, central, and temporal electrodes compared to the occipital and parietal areas.

#### ROC analysis

Slopes discriminated between the unmedicated patients and controls when measured during N2 sleep in all areas, during N3 sleep – in the frontal and temporal areas, and during N1 sleep – in the parietal and occipital areas (AUC=0.68–0.77, all p-values<0.02). Slopes discriminated between the medicated patients and controls when measured during N1, N2, N3, and REM sleep in all areas (AUC=0.62–0.79, all p-values<0.02, Table 1).

### 3.2. Time effect

Within the unmedicated patients, the two-way ANOVA revealed a main effect of the area (F=95.27, p<0.001) and time (early vs late sleep, F=8.50, p=0.006) on the slopes of the N2 epochs with the time-area interaction (F=3.88, p=0.011). The *post hoc* analysis revealed that unmedicated patients showed flatter slopes during late (87 epochs) compared to early (70 epochs) N2 sleep with moderate effect sizes in all areas (p-values=0.002–0.024, Cohen’s d-values=0.42–0.55). Patients in the medicated state and controls showed comparable slopes during late and early N2 sleep in all areas (Fig.2).

**Figure 2.**
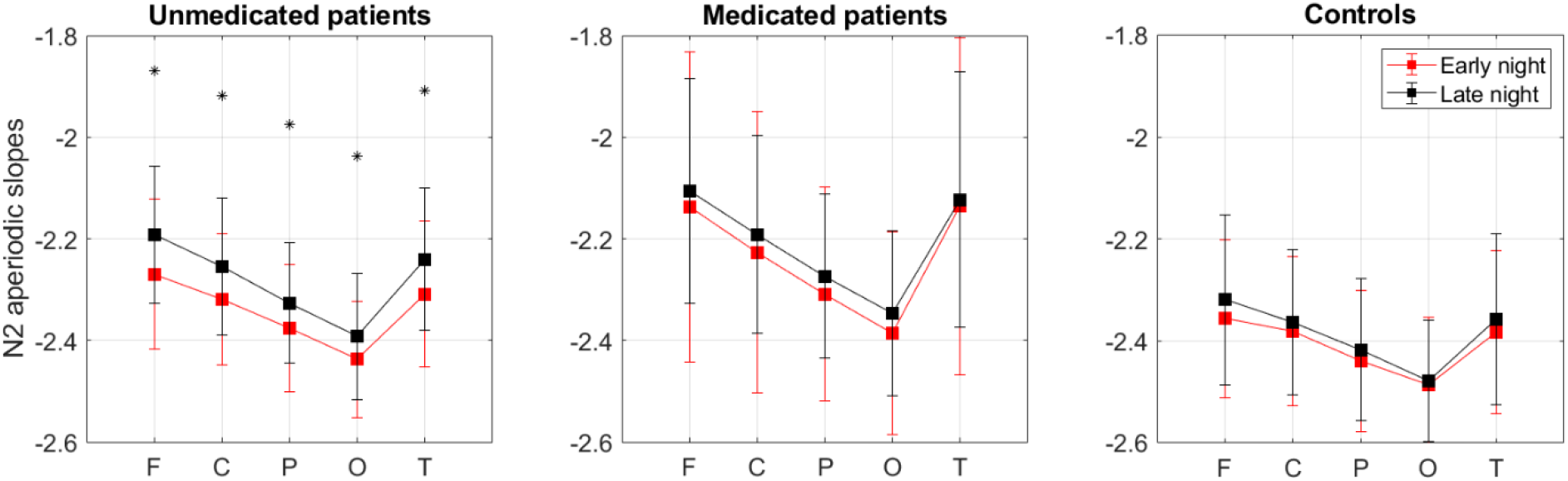
Early vs late N2 aperiodic slopes. The means of the slopes of the broadband (0.2–48 Hz) aperiodic power component of the early (0.5-2 h of sleep) vs late (4.5-6 h of sleep) N2 epochs averaged over each area of interest in each study group separately. Patients in the unmedicated state (left, n=38) show flatter (more positive values) slopes during late (black, 87 epochs) compared to early (red, 70 epochs) N2 sleep in all areas. Patients in the medicated state (middle, n=38) and controls (right, n=38) show comparable slopes during early and late N2 in all areas. F – frontal, C – central, P – parietal, O – occipital, T – temporal electrodes, asterisks mark significant p-values corrected for multiple comparisons, N2 – non-rapid eye movement sleep stage 2.

### 3.3. Correlations

For none of the full-length sleep stages (i.e., non-differentiated by early vs late sleep epochs) at baseline or 7d, the slopes did not correlate with depression severity (HAM-D) at baseline or 7d. After the stratification of the N2 slopes by early vs late epochs, we found that frontal aperiodic slopes of the late but not early N2 epochs at 0d (unmedicated state) negatively correlated with depression severity at 7d (medicated state, Fig.3). There were no correlations between 0d early/late N2 slopes and 0d HAM-D or 7d early/late N2 slopes and 7d HAM-D scores.

**Figure 3.**
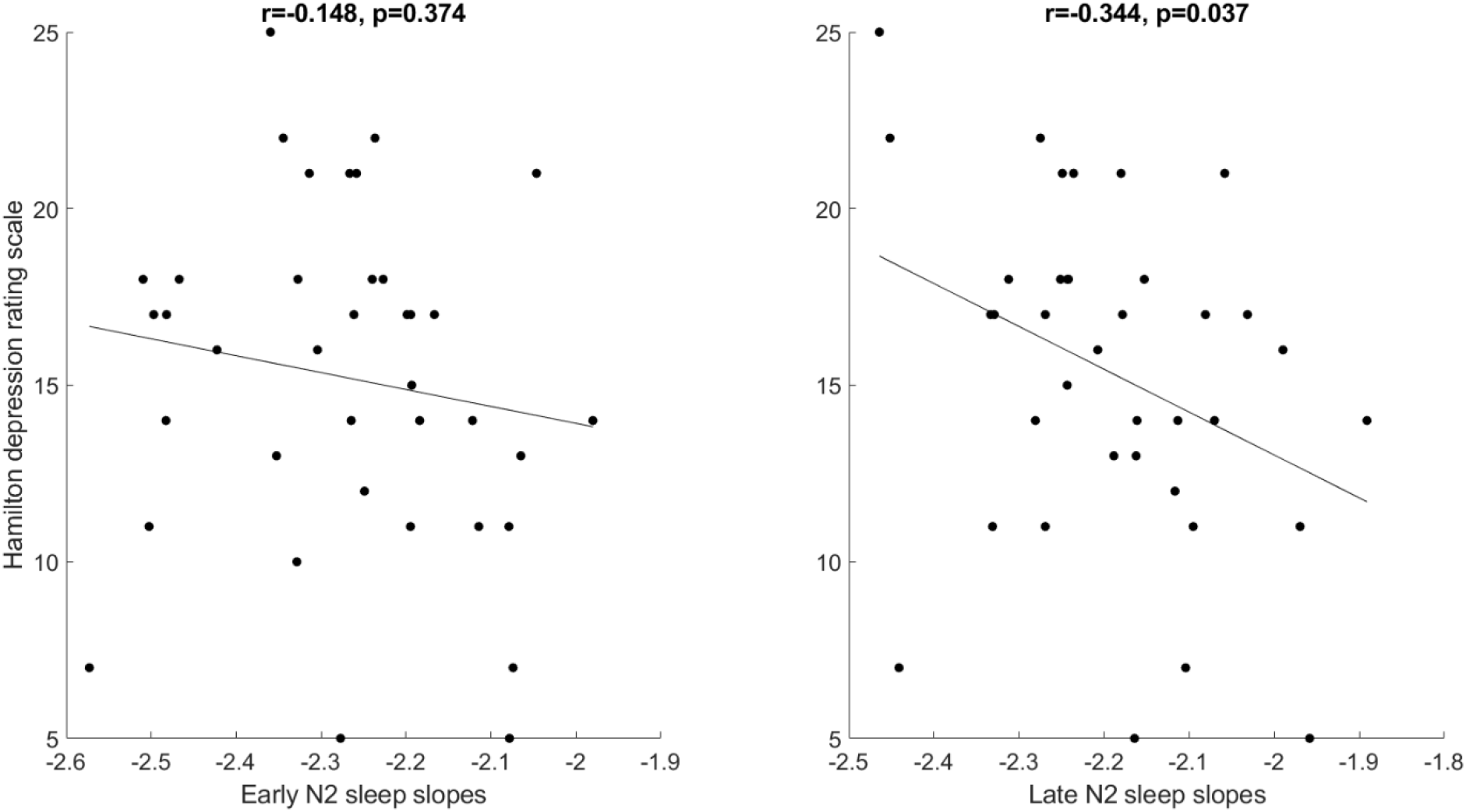
Early and late aperiodic slopes vs depression severity. The slopes of the broadband (0.2–48 Hz) aperiodic power component averaged over early **(left)** or late **(right)** N2 epochs over frontal areas vs the HAM-D scores (depression severity) in unmedicated patients. The slopes of late but not early N2 sleep at 0d negatively correlated with depression severity at 7d, N2 – non-rapid eye movement sleep stage 2, r – Pearson’s correlation coefficients, n=38.

### 3.4. Medication effect

#### REM-suppressive vs REM-non-suppressive

The three-way ANCOVA revealed area-group (F=3.2, p=0.03) and area-stage (F=2.4, p=0.05) interactions. Likewise, there was a strong main effect of the “sleep stage” (F=81.2, p<0.001) and “group” (F=35.7, p<0.001) on the slopes. The two-way ANCOVA revealed a main effect of the group on the slopes measured during the N1 (F=13.9, p=0.001), N2 (F=42.8, p<0.001), and N3 (F=21.9, p<0.001) stages with group-area interactions and during REM sleep (F=10.4, p=0.003) – without a group-area interaction.

The *post hoc* analysis revealed that the patients who took REM-suppressive antidepressants showed flatter slopes during N1, N2, N3, and REM sleep in all areas with large to very large effect sizes (d-values=0.85–1.97) compared to the patients who took REM non-suppressive drugs. During non-REM sleep, the difference between the groups was greater in the frontal and temporal electrodes compared to other areas. Slopes of the wake epochs were comparable (Fig.4, Table S4.2, See also Supplementary Material S4).

**Figure 4.**
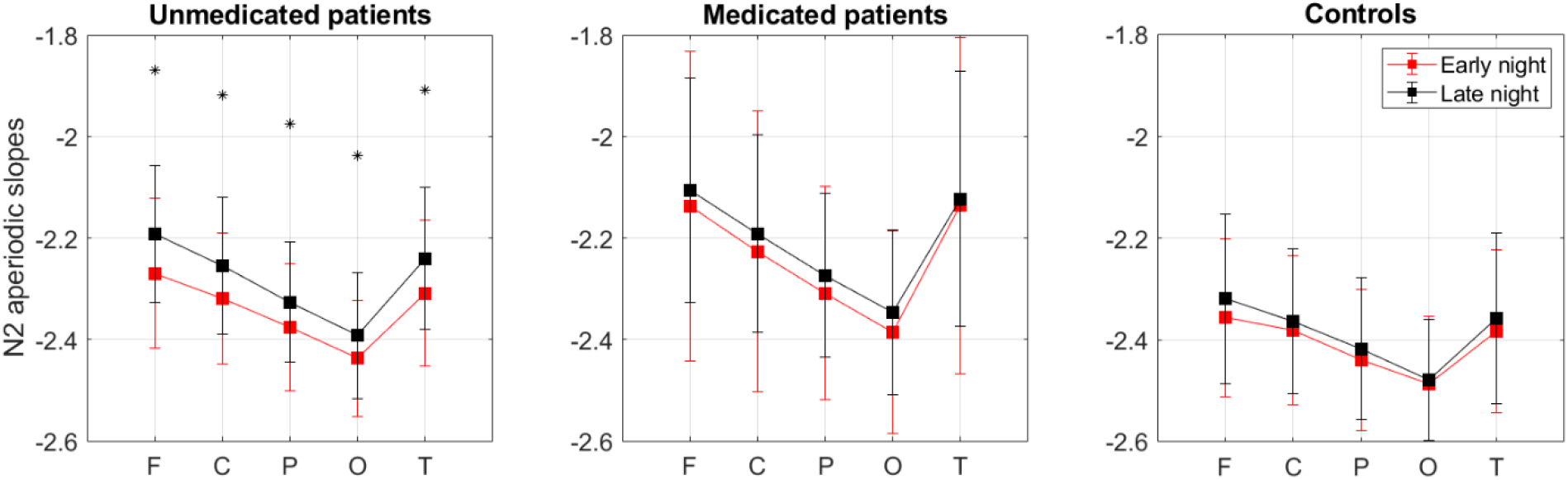
Effect of REM-suppressive medications. The slopes of the aperiodic power component in the 0.2-48Hz frequency band were averaged over each sleep stage over each area of interest. Patients who take REM-suppressive antidepressants for 7d (red, n=21) show flatter slopes (higher values) than patients who take REM non-suppressive antidepressants for 7d (black, n=17) during all sleep stages – but not wake epochs – in all areas. F – frontal, C – central, P – parietal, O – occipital, T – temporal electrodes.

#### Antidepressant classes comparison

The patients who took REM-suppressive SNRIs showed flatter slopes during N2 and REM sleep in all areas, and during the N3 stage – in the frontal, central, and temporal areas with large to huge effect sizes compared to the patients who took non-SNRIs (p-values<0.001–0.026, d-values=0.9– 2.8). These patients also showed flatter slopes compared to the patients who took non-SNRIs REM-suppressive antidepressants (p-values=0.005–0.019, d-values=1.5–1.7), such as SSRIs (p-values=0.009–0.028, d-values=1.3–1.8) during REM sleep in all areas with very large effect sizes. However, these findings did not pass the correction for multiple comparisons. Aperiodic activity was comparable among patients who took SSRIs, TCAs, or NDRIs compared to their pooled controls (Table S4.2, See also Supplementary Material S4).

### 3.4. Variability of slopes

#### Unmedicated patients vs controls

The three-way ANCOVA revealed a main effect of the sleep stage (F=22.4, p<0.001) and area (F=25.2, p<0.001) on the variability of slopes without interactions. The two-way ANCOVA revealed a marginally significant main effect of the group (without a group-area interaction) on the variability of slopes during the N2 (F=6.3, the obtained p=0.015 > the corrected threshold p=0.010), N3 (F=6.2, p=0.015 < the corrected threshold p=0.020) and REM (F=4.7, the obtained p=0.033 > the corrected threshold p=0.030) stages.

The *post hoc* analysis revealed that unmedicated patients showed greater variability of slopes during N2 and N3 sleep in all areas, and during REM sleep – in the central, parietal, and occipital areas with moderate effect sizes compared to controls. Variability of slopes of the wake and N1 epochs was comparable (Fig.1, Table 2).

**Table 2:** Intra-individual variability of slopes.

| Group |  | Unmedicated MDD |  |  |  |  |  | 7d medicated MDD |  |  |  |  |  | HC |  |  |  |  |  |
| --- | --- | --- | --- | --- | --- | --- | --- | --- | --- | --- | --- | --- | --- | --- | --- | --- | --- | --- | --- |
| Mean | Area/Stage | F | C | P | O | T |  | F | C | P | O | T |  | F | C | P | O | T |  |
|  | Wake | 0.45 | 0.43 | 0.36 | 0.35 | 0.44 |  | 0.45 | 0.43 | 0.36 | 0.35 | 0.44 |  | 0.46 | 0.44 | 0.37 | 0.36 | 0.46 |  |
|  | N1 | 0.31 | 0.30 | 0.27 | 0.26 | 0.34 |  | 0.31 | 0.30 | 0.27 | 0.26 | 0.34 |  | 0.30 | 0.29 | 0.26 | 0.25 | 0.35 |  |
|  | N2 | 0.27 | 0.25 | 0.23 | 0.22 | 0.28 |  | 0.27 | 0.25 | 0.23 | 0.22 | 0.28 |  | 0.25 | 0.23 | 0.21 | 0.12 | 0.26 |  |
|  | N3 | 0.22 | 0.20 | 0.18 | 0.17 | 0.23 |  | 0.22 | 0.20 | 0.18 | 0.17 | 0.23 |  | 0.19 | 0.16 | 0.15 | 0.14 | 0.19 |  |
|  | REM | 0.22 | 0.18 | 0.15 | 0.14 | 0.20 |  | 0.22 | 0.18 | 0.15 | 0.14 | 0.20 |  | 0.19 | 0.14 | 0.12 | 0.12 | 0.17 |  |
| Comparison |  | Unmedicated MDD-HC |  |  |  |  |  | 7d medicated MDD-HC |  |  |  |  |  | Unmedicated-medicated MDD |  |  |  |  |  |
| Effect size | Area/Stage | F | C | P | O | T |  | F | C | P | O | T |  | F | C | P | O | T |  |
|  | Wake | -0.06 | -0.11 | -0.10 | -0.13 | -0.15 |  | -0.03 | -0.23 | -0.16 | -0.08 | -0.24 |  | -0.02 | 0.09 | 0.04 | -0.05 | 0.06 |  |
|  | N1 | 0.04 | 0.19 | 0.09 | 0.10 | -0.05 |  | 0.15 | 0.19 | -0.13 | -0.03 | 0.14 |  | -0.08 | 0.03 | 0.15 | 0.09 | -0.18 |  |
|  | N2 | 0.46 | 0.66 | 0.59 | 0.57 | 0.49 |  | 0.72 | 0.85 | 0.70 | 0.77 | 1.08 |  | -0.21 | -0.15 | -0.02 | -0.06 | -0.37 |  |
|  | N3 | 0.33 | 0.66 | 0.60 | 0.48 | 0.56 |  | 0.68 | 0.74 | 0.71 | 0.59 | 0.68 |  | -0.31 | -0.12 | -0.09 | -0.08 | -0.13 |  |
|  | REM | 0.42 | 0.60 | 0.53 | 0.51 | 0.38 |  | 0.53 | 0.50 | 0.62 | 0.63 | 0.67 |  | 0.02 | 0.15 | 0.06 | 0.06 | -0.12 |  |
| Comparison |  | Unmedicated MDD-HC |  |  |  |  |  | 7d medicated MDD-HC |  |  |  |  |  | Unmedicated-medicated MDD |  |  |  |  |  |
| Factor |  | Group |  | Area |  | Interaction |  | Group |  | Area |  | Interaction |  | Group |  | Area |  | Interaction |  |
|  |  | F | p | F | p | F | P | F | p | F | p | F | p | F | p | F | p | F | p |
| ANOVA | Wake | 0.3 | 0.58 | 4.1 | 0.01 | 0.1 | 0.73 | 0.5 | 0.46 | 3.1 | 0.02 | 0.6 | 0.61 | 0.0 | 0.89 | 9.8 | 0.00 | 0.3 | 0.83 |
|  | N1 | 0.1 | 0.75 | 15.8 | 0.00 | 0.7 | 0.51 | 1.2 | 0.66 | 12.8 | 0.00 | 1.0 | 0.39 | 0.0 | 0.98 | 12.9 | 0.00 | 1.7 | 0.17 |
|  | N2 | 6.3 | 0.02 | 23.2 | 0.00 | 0.7 | 0.61 | 16.8 | 0.00 | 14.0 | 0.00 | 5.0 | 0.04 | 1.3 | 0.26 | 17.2 | 0.00 | 2.9 | 0.05 |
|  | N3 | 6.2 | 0.02 | 5.3 | 0.00 | 1.9 | 0.14 | 10.4 | 0.00 | 8.9 | 0.00 | 2.6 | 0.07 | 0.4 | 0.52 | 6.4 | 0.00 | 1.3 | 0.26 |
|  | REM | 4.7 | 0.03 | 15.1 | 0.00 | 1.2 | 0.22 | 8.2 | 0.01 | 15.6 | 0.00 | 1.8 | 0.15 | 0.0 | 0.86 | 18.3 | 0.00 | 2.8 | 0.05 |

***Bold font indicates statistically significant p-values after the correction for multiple comparisons, MDD – major depressive disorder, HC – healthy controls, F – frontal, C – central, P – parietal, O – occipital, T – temporal electrodes, REM – rapid eye movement sleep, N – non-rapid eye movement sleep.***

#### Medicated patients vs controls

The three-way ANCOVA revealed a stage-group interaction (F=3.4, p=0.01). The two-way ANCOVA revealed a main effect of the group without a group-area interaction on the variability of slopes during N3 (F=10.4, the obtained p=0.002 < the corrected threshold p=0.020) and REM (F=8.2, the obtained p=0.006 < the corrected threshold p=0.030) sleep. In addition, there was a main effect of the group with a group-area interaction during the N2 stage (F=16.8, p<0.001).

The *post hoc* analysis revealed that medicated patients showed greater variability of slopes during the N2, N3, and REM stages in all areas with moderate to large effect sizes compared to controls. During the N2 stage, the difference was smaller in the parietal and occipital electrodes compared to the frontal, central, and temporal areas. Variability of slopes of the wake and N1 epochs was comparable (Fig.1, Table 2).

These findings were replicated using two independently collected datasets of short and long-term medicated patients (Supplementary Material S4).

#### Unmedicated vs medicated states

Unmedicated and medicated states showed comparable variability of slopes during all stages in all areas (Fig.1, Table 2).

The reported in the Results findings were specific to sleep and were not observed during the morning resting state (reported in Supplementary Material 2).

## 4. Discussion

To the best of our knowledge, this is the first study that examines sleep-related aperiodic activity in MDD and its relationships with the time of sleep, depression severity, and responsivity to antidepressant treatment. We found that unmedicated patients show flatter aperiodic slopes during non-REM sleep with increased variability during both non-REM and REM sleep compared to controls. Within unmedicated patients, aperiodic slopes are flatter during late compared to early non-REM sleep. Flatter frontal slopes of late non-REM sleep in the unmedicated state are further associated with lower depression severity at 7d after the commencement of antidepressant treatment. Patients in the medicated state show flatter aperiodic slopes compared to the own unmedicated state and healthy controls during both non-REM and REM sleep. We replicated several of our findings in two independently collected datasets of medicated patients.

The statistical analyses showed that all these effects are significant and rather strong, but what is the neurophysiological meaning of slope flattering? The functional significance of aperiodic dynamics is still a mystery with several interpretations suggested so far. For example, aperiodic activity can manifest in the overall firing rate of cortical neurons (9, 14, 22) as measured by local field potentials or EEG. When many neurons fire relatively simultaneously, the power spectrum will decay faster, being relatively stronger in low frequencies and relatively weaker in the higher ones. Mathematically, this will be expressed by a more negative (steeper) slope, which, in turn, reflects a higher power-law exponent. A steeper slope can signify redundancy (8), excessive or insufficient propagation of the signal (23), or increased dendritic filtering (22). When neurons fire relatively asynchronously, the spectral power is shifted towards higher frequencies and its slope is flatter, reflecting reduced temporal autocorrelations (6), high entropy rate of cortical systems (24), or a noisier neural background (25, 26). Following this, flatter aperiodic slopes observed here in unmedicated and medicated MDD patients may reflect noisier neural background activity, which in turn can adversely affect sleep and its restorative function.

Adding to this, pharmacological, physiological, and computational studies linked aperiodic activity to the balance between excitatory and inhibitory currents in the brain (12, 21). Specifically, Gao et al. (12) showed that the aperiodic slope reliably tracked the induction and the recovery from propofol-induced anesthesia in rats and macaques. Similarly, in humans, inhibition was boosted by the propofol administration, and the slope became steeper when inhibition increased (21). The right balance between neural excitation and inhibition is crucial for optimal signal formation and transmission, synaptic plasticity, neuronal growth, and pruning and, thus, enables flexible behavior and cognition (13). Correspondingly, any perturbations in the E/I balance may lead to brain disease (13). For example, autism has been linked to a high E/I ratio caused by hypoactive receptors for the brain’s principal inhibitory neurotransmitter GABA (13). Schizophrenia has been associated with a low E/I ratio caused by hypoactive receptors for the excitatory neurotransmitter glutamate (13). Interestingly, patients with schizophrenia show steeper aperiodic slopes compared to controls during rest (27), while patients with Tourette syndrome (with its dopaminergic and GABAergic alterations) present flatter aperiodic slopes during sensorimotor processing, reflecting increased neural noise (28). Analogously, depression might be associated with E/I perturbations due to its cholinergic-monoaminergic (14, 15), glutamatergic (16), and/or GABAergic imbalance.

Indeed, it has been suggested that GABAergic deficit may play a central role in the etiology of MDD, especially in melancholic (17) and treatment-resistant types of depression (18). Moreover, targeting the E/I imbalance in depression via enhancing the GABAergic system with antidepressant therapies may contribute to a greater remission rate and reduce the risk of relapse (29). At the cellular level, changes in GABAergic interneurons affect the regulation of excitatory signals from and onto pyramidal neurons (30), the primary contributors to the EEG signal. Following this literature, flatter aperiodic slopes with increased variability observed here in unmedicated and medicated patients may reflect a shift in the E/I ratio in favor of excitation with higher background neural noise due to cellular alterations of the GABAergic, glutamatergic, and cholinergic-monoaminergic systems. Behaviorally, this can turn sleep into a more unstable and easy-to-disrupt state than it should be. Noisy neural activity may cause a shift from external to internal (own) mental contents in awareness, which is typical of MDD (30). Such a shift could result in increased inner tension, self-focus, and rumination, probably causing difficulties to relax and initiate or maintain sleep (30). Impaired sleep could in turn lead to chronic stress, which affects GABAergic transmission, which is vital for stress control (17). Importantly, aperiodic alterations observed in this study were specific to sleep and were not observed during the wake epochs of sleep or morning resting state (Supplementary Material 2).

Interestingly, patients showed flatter aperiodic slopes during late compared to early non-REM sleep. This effect was observed only in the unmedicated state, suggesting that it can be partially attributable to circadian disturbances intrinsic to MDD. Flatter frontal slopes during late non-REM sleep in the unmedicated state were further associated with lower depression severity at 7d after the commencement of antidepressant treatment. In other words, the patients who were somehow able to “flatten” their aperiodic activity towards morning showed a better later response to antidepressant treatment (as reflected by lower HAM-D scores measured 7d after the commencement of antidepressant treatment). Keeping in mind that healthy participants show steeper – and not flatter – slopes compared to unmedicated patients, this finding looks very intriguing. A possible interpretation is that it reflects individual compensatory abilities of patients, for example, an effort to compensate for the disorder, using different (compared to healthy controls) neural pathways as a response to excitation-to-inhibition imbalance. Nevertheless, it should be stressed that this sub-analysis was exploratory and future studies are needed to confirm it and search for the mechanisms underlying the reported findings.

Of special interest was the effect of antidepressants: we found that during all sleep stages, medicated patients showed flatter slopes compared to controls. We replicated this association using two independently collected datasets of short and long-term medicated patients (Supplementary Material 4). In addition, we found that the medicated state showed flatter slopes compared to the own unmedicated state. In line with our findings, a recent study in healthy females has reported that one week of intake of the SSRI escitalopram induces a flattening of aperiodic slopes during rest in favor of excitation (31). Assuming that flatter aperiodic slopes reflect noisier neural background and, therefore, less efficient sleep, our record confirms the previous studies stating that some antidepressants (for example, such SNRI as venlafaxine) may disrupt sleep due to their activating effects (32). Given that a deeper understanding of the antidepressants’ effects on sleep is crucial for the successful treatment, future large-scale longitudinal research is required to confirm whether aperiodic activity can serve as a marker for predicting individual cortical responsivity to different antidepressants.

In this study, we also found a strong within-subject topographical effect. This is in line with a recent study that reports spatial heterogeneity of aperiodic activity such that the slopes become gradually flatter as they move anteriorly (6). Here, the difference between the groups was more pronounced in the frontal and temporal electrodes, possibly reflecting molecular alterations at the cellular level. This agrees with the finding that GABAergic deficits seen in MDD are specific to prefrontal somatostatin-expressing and, to a lesser extent, hippocampal parvalbumine-positive interneurons (29). One should keep in mind, however, that due to the volume conduction, electrodes placed over a given region can also capture EEG activity from other areas (34).

Finally, it is important to note that in our previous study, we could not detect differences in the total (i.e., non-differentiated to its components) spectral power between unmedicated patients and controls (5). Taken together our recent and current reports demonstrate that in unmedicated patients with MDD, impaired sleep might be related to altered aperiodic activity specifically. Besides its scientific importance, this finding is also essential from the methodological point of view as it confirms the importance of a recent recommendation to differentiate the total spectral power to its components in order “to avoid misrepresentation and misinterpretation of the data” (6,7). Of note, given that aperiodic activity can be contaminated by residual slow-wave oscillatory activity, in the Supplementary Material 1 we report the aperiodic analysis performed in the 2-20 Hz (to exclude low oscillatory activity at all) and 30-48 Hz bands (to analyze high-frequency activity separately) (12, 21). These supplementary analyses confirm the broadband analysis reported in the main text.

This study is not without limitations. First, one should keep in mind that the diagnosis of depression is subjective, and subtypes of depression likely exist even though they have not been systematically distinguished. Likewise, the stratification of the patients by antidepressant classes performed here was not clean enough as the patients used different antidepressants. Second, this research is correlational and precludes causal relations between the brain and disorder. Despite the aforementioned limitations, this study provides a new facet of the neurobiology of sleep in depression and the pharmacological effect of antidepressants.

In conclusion, our findings suggest that flatter aperiodic slopes with increased variability represent a new disease-relevant feature of sleep in MDD, which may reflect unstable, noisy neural activity due to a shift of the excitation-to-inhibition ratio in favor of excitation. In the future, these findings may lead to the development of a biomarker for personalized disease monitoring and therapy.

## Data Availability

All data produced in the present study are available upon reasonable request to the authors

## Acknowledgments

We would like to thank all the participants for participating in this study. We would like to thank Sofia Tzioridou and Dr. Sarah Schoch for their helpful suggestions.

## Contributors

MD and MZ designed the study. YR analyzed the data and wrote the manuscript. All authors contributed to, reviewed, and approved the final draft of the paper. All authors had full access to all the data in the study and had final responsibility for the decision to submit for publication.

## Data availability

All data produced in the present study are available upon reasonable request to the authors.

## Abbreviations

AUC: area under the curve
E/I: excitation-to-inhibition
HAM-D: Hamilton depression rating scale
MDD: major depressive disorder
NaSSA: noradrenergic and specific serotonergic antidepressants
NDRI: norepinephrine-dopamine reuptake inhibitor
REM: rapid eye movement
SNRI: serotonin-norepinephrine reuptake inhibitors
SSRI: selective serotonin reuptake inhibitors
TCA: tricyclic antidepressants

