## Supplementary Material S1 for "Increased aperiodic neural activity during sleep in major depressive disorder"

Martin Dresler [1,2]

[1]Radboud University, Donders Institute for Brain, Cognition and Behavior, Nijmegen, Netherlands,
[2]Radboud University Medical Centre, Department of Cognitive Neuroscience, Nijmegen, Netherlands,
[3]Netherlands Institute for Neuroscience, Department of Sleep and Cognition, Amsterdam, Netherlands,
[4]Max Planck Institute of Psychiatry, Munich, Germany, [5]Klinikum Ingolstadt, Centre of Mental Health, Ingolstadt, Germany

### Supplementary Material 1

#### Low and high band aperiodic power

In the Main Text, we analyzed the broadband (0.2-48Hz) signal. Here, we further report low (2-20Hz) and high (30-48Hz) frequency bands analyses. The low-band analysis was added to this work to control for a possible distortion of the linear fit of the aperiodic component by excluding low frequencies with residual (failed to be excluded by the IRASA[1] algorithm) strong oscillatory activity as well as the so-called "knees", specific bends seen on the power spectrum in log-log space around 1-2Hz and 20Hz (2). We added the high-band analysis inspired by the literature reporting that this band facilitates the reliable discrimination between wakefulness and REM sleep

[1] IRASA – Irregularly Resampled Auto-Spectral Analysis (1).

(3). The analysis was limited to 48 Hz due to line noise in recordings (50Hz in Europe) and broadband muscle artifacts (3).

#### Results

##### S1.1. Means of low-band slopes

*Unmedicated patients vs controls*

The three-way ANCOVA revealed stage-group (F=4.3, p=0.01) and area-stage (F=9.3, p<0.001) interactions. The two-way ANCOVA revealed a main effect of the group with a group-area interaction on the slopes measured during the N2 (F=9.6, the obtained p=0.003 < the corrected threshold p=0.010) and N3 (F=8.0, the obtained p=0.006 < the corrected threshold p=0.020) stages.

The *post hoc* analysis revealed that unmedicated patients showed flatter slopes during the N2 and N3 stages in all areas with moderate effect sizes compared to controls. The difference between the groups was greater in the frontal electrodes compared to other areas. Slopes during the wake, N1, and REM epochs were comparable (Fig. S1.1, Table S1.1).

*7d medicated patients vs controls*

The three-way ANCOVA revealed stage-group (F=8.7, p<0.001) and area-stage (F=5.8, p<0.001) interactions. The two-way ANCOVA revealed an effect of the group on the slopes measured during the N2 and N3 stages (p-values<0.001) with an area-group interaction and during the N1 (F=7.7, p=0.006) and REM (F=7.5, p=0.008) stages without interactions.

The *post hoc* analysis revealed that medicated patients showed flatter slopes during N1, N2, and N3 sleep in all areas with moderate to large effect sizes compared to controls. During N2 and N3 sleep, the difference between the groups was greater in the frontal and temporal electrodes compared to other areas. During REM sleep, medicated patients showed flatter slopes in the central, parietal, occipital, and temporal areas with moderate effect sizes compared to controls. Slopes during the wake epochs were comparable (Fig. S1.1, Table S1.1).

*Unmedicated vs medicated states*

The three-way ANOVA revealed a main effect of the group ($F=8.8$, $p=0.005$) with an area-stage interaction ($F=25.4$, $p<0.001$). The two-way ANCOVA revealed a main effect with area-state interactions for the N1 ($F=7.8$, $p=0.008$), N2 ($F=6.9$, $p=0.012$), N3 ($F=7.3$, $p=0.009$), and REM ($F=11.3$, $p=0.002$) stages.

The *post hoc* analysis revealed that patients showed flatter slopes when medicated than when unmedicated during the N2 and N3 stages in all areas, and during the N1 and REM stages – in the central, parietal, occipital, and temporal areas with moderate effect sizes. During N1 and N2 sleep, the difference between the states was greater in the temporal area, during N3 sleep – in the central and temporal areas, and during REM sleep – in the occipital electrodes compared to other areas. Slopes during the wake epochs were comparable (Fig. S1.1, Table S1.1).

**Table S1.1: Slope means**

| Mean | Groups | Unmedicated MDD | | | | | 7d medicated MDD | | | | | HC | | | | |
|---|---|---|---|---|---|---|---|---|---|---|---|---|---|---|---|---|
| Low band | Area/ Stage | F | C | P | O | T | F | C | P | O | T | F | C | P | O | T |
| | Wake | -1.30 | -1.23 | -1.20 | -1.22 | -1.02 | -1.36 | -1.18 | -1.23 | -1.22 | -0.95 | -1.25 | -1.12 | -1.132 | -1.148 | -0.90 |
| | N1 | -1.63 | -1.59 | -1.63 | -1.72 | -1.58 | -1.58 | -1.52 | -1.58 | -1.67 | -1.47 | -1.73 | -1.65 | -1.690 | -1.781 | -1.65 |
| | N2 | -1.92 | -1.93 | -1.97 | -2.06 | -1.93 | -1.86 | -1.87 | -1.93 | -2.03 | -1.85 | -2.08 | -2.05 | -2.074 | -2.161 | -2.06 |
| | N3 | -2.53 | -2.53 | -2.57 | -2.63 | -2.57 | -2.45 | -2.47 | -2.19 | -2.60 | -2.48 | -2.69 | -2.65 | -2.685 | -2.740 | -2.71 |
| | REM | -1.77 | -1.68 | -1.70 | -1.73 | -1.72 | -1.75 | -1.61 | -1.62 | -1.64 | -1.64 | -1.85 | -1.74 | -1.763 | -1.796 | -1.78 |
| High band | Area/ Stage | F | C | P | O | T | F | C | P | O | T | F | C | P | O | T |
| | Wake | -1.21 | -1.38 | -1.75 | -1.92 | -1.24 | -1.06 | -1.22 | -1.52 | -1.64 | -1.00 | -1.07 | -1.27 | -1.59 | -1.72 | -1.06 |
| | N1 | -2.46 | -2.88 | -3.08 | -3.15 | -2.67 | -2.28 | -2.68 | -2.91 | -2.98 | -2.39 | -2.49 | -2.96 | -3.13 | -3.19 | -2.72 |
| | N2 | -2.69 | -3.16 | -3.32 | -3.39 | -3.02 | -2.43 | -2.89 | -3.07 | -3.16 | -2.64 | -2.72 | -3.24 | -3.34 | -3.41 | -3.05 |
| | N3 | -2.58 | -3.09 | -3.22 | -3.28 | -2.91 | -2.32 | -2.80 | -2.95 | -3.05 | -2.56 | -2.59 | -3.16 | -3.20 | -3.26 | -2.93 |
| | REM | -2.73 | -2.108 | -3.26 | -3.37 | -3.145 | -2.54 | -2.82 | -3.07 | -3.18 | -2.90 | -2.68 | -2.95 | -3.17 | -3.29 | -3.05 |
| **Effect size** | **Groups** | **Unmedicated MDD-HC** | | | | | **Medicated MDD-HC** | | | | | **Unmedicated-medicated MDD** | | | | |
| Low band | Area/ Stage | F | C | P | O | T | F | C | P | O | T | F | C | P | O | T |
| | Wake | -0.12 | -0.28 | -0.18 | -0.22 | -0.30 | -0.21 | -0.14 | -0.29 | -0.24 | -0.15 | 0.11 | -0.13 | 0.12 | -0.01 | -0.16 |
| | N1 | 0.40 | 0.22 | 0.26 | 0.27 | 0.29 | **0.63** | **0.55** | **0.49** | **0.47** | **0.70** | -0.30 | **-0.51** | **-0.37** | -0.32 | **-0.57** |
| | N2 | **0.88** | **0.70** | **0.63** | **0.58** | **0.68** | **1.01** | **0.88** | **0.77** | **0.70** | **0.96** | **-0.38** | **-0.40** | **-0.36** | -0.32 | **-0.53** |
| | N3 | **0.70** | **0.57** | **0.55** | **0.52** | **0.63** | **0.95** | **0.85** | **0.78** | **0.74** | **0.94** | **-0.36** | **-0.40** | **-0.34** | **-0.28** | **-0.40** |
| | REM | 0.33 | 0.235 | 0.258 | 0.27 | 0.25 | 0.42 | **0.56** | **0.60** | **0.62** | **0.53** | -0.13 | **-0.53** | **-0.67** | **-0.74** | **-0.53** |
| High band | Area/ Stage | F | C | P | O | T | F | C | P | O | T | F | C | P | O | T |
| | Wake | -0.21 | -0.15 | -0.24 | -0.32 | -0.29 | 0.02 | 0.07 | 0.10 | 0.12 | 0.11 | -0.21 | -0.19 | -0.30 | **-0.38** | -0.37 |
| | N1 | 0.07 | 0.22 | 0.14 | 0.12 | 0.10 | **0.52** | **0.62** | **0.56** | **0.58** | **0.68** | **-0.42** | **-0.40** | **-0.42** | **-0.45** | **-0.49** |

| | | | | | | | | | | | | | | | | |
|---|---|---|---|---|---|---|---|---|---|---|---|---|---|---|---|---|
| | N2 | 0.10 | 0.30 | 0.10 | 0.07 | 0.09 | **0.73** | **0.84** | **0.73** | **0.73** | **0.92** | **-0.57** | **-0.54** | **-0.54** | **-0.57** | **-0.71** |
| | N3 | 0.03 | 0.22 | -0.06 | -0.08 | 0.09 | **0.66** | **0.81** | **0.65** | **0.60** | **0.75** | **-0.55** | **-0.57** | **-0.58** | **-0.53** | **-0.65** |
| | REM | -0.11 | -0.11 | -0.19 | -0.22 | -0.17 | 0.34 | 0.29 | 0.25 | 0.28 | 0.35 | **-0.57** | **-0.64** | **-0.73** | **-0.78** | **-0.76** |

***Bold*** *font indicates statistically significant p-values after the correction for multiple comparisons, MDD – major depressive disorder, HC – healthy controls, F – frontal, C – central, P – parietal, O – occipital, T – temporal electrodes, REM – rapid eye movement sleep, N – non–rapid eye movement sleep.*

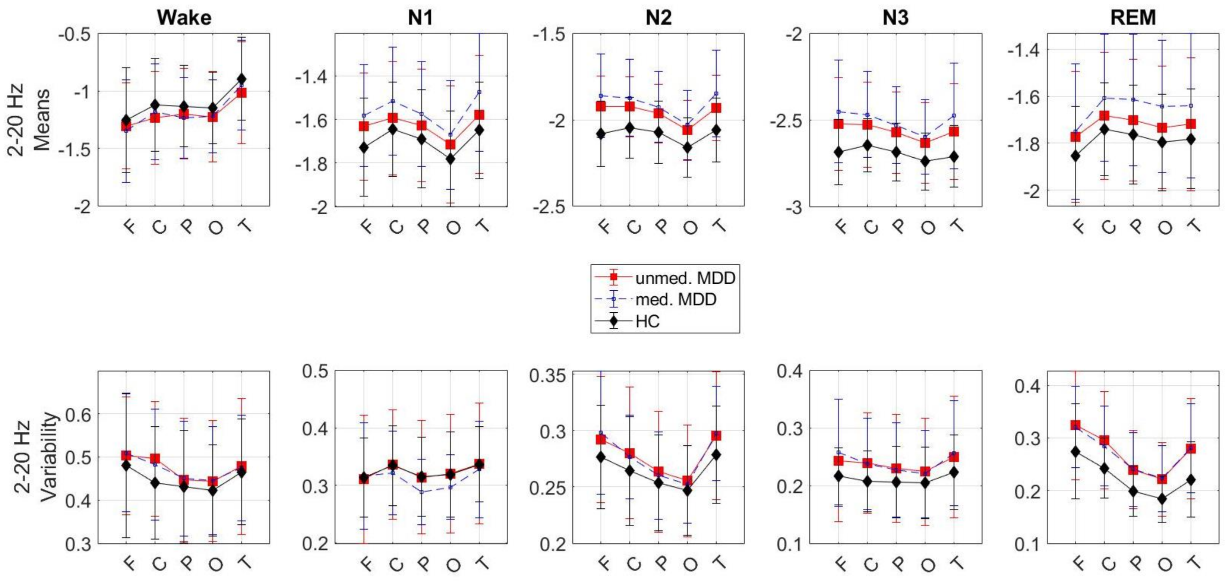


**Figure S1.1. Means and variability of the low-band slopes.** Means and intra-individual variability of the slopes of the aperiodic power component in the low (2-20Hz) frequency band over each sleep stage over each area of interest in each study group. **Top:** Unmedicated patients (red) show flatter (more positive values) slopes during N2 and N3 stages compared to controls (black) in all areas. The flattest slopes are observed in the frontal area. 7d medicated patients (blue) show flatter slopes compared to the own unmedicated state (red) and compared to controls (black) during all sleep stages in all areas. **Bottom:** Patients in both unmedicated and 7d medicated states show greater intra-individual variability of slopes during REM sleep compared to controls. MDD – 38 major depressive disorder patients, unmed. – unmedicated, med. - 7d medicated, HC – 38 healthy controls, F – frontal, C – central, P – parietal, O – occipital, T – temporal electrodes.

#### S1.2. Variability of low-band slopes

*Unmedicated patients vs controls*

The three-way ANCOVA revealed a strong main effect of the sleep stage ($F=19.2$, $p<0.001$), area ($F=7.6$, $p<0.001$), and the group ($F=4.8$, $p=0.03$) on the variability of slopes without interactions. The two-way ANCOVA revealed a main effect of the group without a group-area interaction on the variability of slopes during REM sleep ($F=9.1$, the obtained $p=0.004$ < the corrected threshold $p=0.010$). The *post hoc* analysis revealed that unmedicated patients showed greater variability of slopes during REM sleep in all areas with moderate effect sizes compared to controls. Variability of slopes during the wake, N1, N2, and N3 epochs was comparable (Fig. S1.1, Table S1.2).

*7d medicated patients vs controls*

The three-way ANCOVA revealed a strong main effect of the sleep stage ($F=13.5$, $p<0.001$), area ($F=8.1$, $p<0.001$) and group ($F=7.7$, $p=0.007$) on the variability of slopes without interactions. The two-way ANCOVA revealed a main effect of the group on the variability of slopes without a group-area interaction during REM sleep ($F=10.7$, the obtained $p=0.002$ < the corrected threshold $p=0.010$). The *post hoc* analysis revealed that medicated patients showed greater variability of slopes during REM sleep in all areas with moderate effect sizes compared to controls. Variability of slopes during the wake, N1, N2, and N3 epochs was comparable (Fig. S1.1, Table S1.2).

**Table S1.2: Intra-individual variability of slopes**

| Std | Group | Unmedicated MDD | | | | | 7d medicated MDD | | | | | HC | | | | |
|---|---|---|---|---|---|---|---|---|---|---|---|---|---|---|---|---|
| Low band | Area/ Stage | F | C | P | O | T | F | C | P | O | T | F | C | P | O | T |
| | Wake | 0.50 | 0.50 | 0.45 | 0.44 | 0.48 | 0.51 | 0.48 | 0.45 | 0.45 | 0.48 | 0.48 | 0.44 | 0.43 | 0.42 | 0.47 |
| | N1 | 0.31 | 0.34 | 0.31 | 0.32 | 0.34 | 0.32 | 0.32 | 0.29 | 0.30 | 0.33 | 0.31 | 0.33 | 0.32 | 0.32 | 0.34 |
| | N2 | 0.29 | 0.28 | 0.26 | 0.26 | 0.305 | 0.30 | 0.28 | 0.26 | 0.25 | 0.30 | 0.28 | 0.26 | 0.25 | 0.25 | 0.28 |
| | N3 | 0.24 | 0.24 | 0.23 | 0.23 | 0.25 | 0.26 | 0.24 | 0.16 | 0.22 | 0.26 | 0.22 | 0.21 | 0.21 | 0.21 | 0.22 |
| | REM | 0.32 | 0.30 | 0.24 | 0.22 | 0.28 | 0.32 | 0.28 | 0.24 | 0.22 | 0.28 | 0.28 | 0.24 | 0.20 | 0.18 | 0.22 |
| High band | Area/ Stage | F | C | P | O | T | F | C | P | O | T | F | C | P | O | T |
| | Wake | 0.89 | 0.99 | 0.92 | 0.88 | 0.87 | 0.88 | 0.98 | 0.94 | 0.90 | 0.85 | 0.92 | 1.05 | 1.00 | 0.95 | 0.91 |
| | N1 | 0.69 | 0.74 | 0.65 | 0.60 | 0.84 | 0.71 | 0.78 | 0.69 | 0.64 | 0.88 | 0.67 | 0.71 | 0.61 | 0.58 | 0.84 |
| | N2 | 0.52 | 0.54 | 0.47 | 0.43 | 0.62 | 0.55 | 0.58 | 0.52 | 0.48 | 0.68 | 0.49 | 0.48 | 0.42 | 0.39 | 0.56 |
| | N3 | 0.45 | 0.44 | 0.38 | 0.35 | 0.51 | 0.45 | 0.47 | 0.33 | 0.40 | 0.51 | 0.44 | 0.39 | 0.34 | 0.32 | 0.44 |
| | REM | 0.47 | 0.57 | 0.52 | 0.47 | 0.53 | 0.46 | 0.54 | 0.49 | 0.45 | 0.55 | 0.46 | 0.55 | 0.49 | 0.43 | 0.51 |
| **Effect size** | **Group** | **Unmedicated MDD-HC** | | | | | **7d medicated MDD-HC** | | | | | **Unmedicated-7d medicated MDD** | | | | |
| Low band | Area/ Stage | F | C | P | O | T | F | C | P | O | T | F | C | P | O | T |
| | Wake | 0.15 | 0.42 | 0.11 | 0.17 | 0.09 | 0.19 | 0.33 | 0.14 | 0.19 | 0.07 | -0.04 | 0.09 | -0.02 | -0.01 | 0.02 |
| | N1 | -0.03 | 0.02 | -0.01 | 0.02 | 0.02 | 0.03 | -0.18 | -0.42 | -0.33 | -0.11 | -0.05 | 0.13 | 0.26 | 0.22 | 0.10 |
| | N2 | 0.30 | 0.30 | 0.21 | 0.19 | 0.34 | 0.43 | 0.29 | 0.17 | 0.15 | 0.45 | -0.09 | 0.06 | 0.06 | 0.06 | -0.03 |
| | N3 | 0.33 | 0.43 | 0.29 | 0.25 | 0.30 | 0.55 | **0.45** | **0.29** | **0.23** | **0.42** | -0.17 | 0.02 | 0.04 | 0.07 | -0.10 |
| | REM | **0.51** | **0.69** | **0.65** | **0.65** | **0.70** | 0.54 | 0.62 | 0.69 | 0.72 | 0.76 | 0.05 | 0.12 | -0.01 | -0.01 | -0.10 |
| High band | Area/ Stage | F | C | P | O | T | F | C | P | O | T | F | C | P | O | T |
| | Wake | -0.10 | -0.21 | -0.30 | -0.27 | -0.12 | -0.13 | -0.22 | -0.21 | -0.19 | -0.19 | 0.03 | 0.02 | -0.06 | -0.06 | 0.08 |
| | N1 | 0.10 | 0.16 | 0.21 | 0.12 | -0.01 | 0.21 | 0.39 | 0.45 | 0.36 | 0.24 | -0.09 | -0.20 | -0.23 | -0.23 | -0.25 |

| | | | | | | | | | | | | | | | | |
|---|---|---|---|---|---|---|---|---|---|---|---|---|---|---|---|---|
| | N2 | 0.23 | 0.46 | 0.50 | 0.39 | 0.39 | 0.43 | **0.69** | **0.78** | **0.74** | **0.80** | -0.19 | -0.24 | -0.29 | -0.32 | -0.39 |
| | N3 | 0.05 | 0.29 | 0.30 | 0.24 | 0.37 | 0.10 | **0.45** | **0.52** | 0.57 | 0.37 | -0.04 | -0.17 | -0.21 | -0.29 | 0.03 |
| | REM | 0.07 | 0.20 | 0.30 | 0.32 | 0.17 | -0.03 | -0.05 | 0.03 | 0.15 | 0.28 | 0.06 | 0.22 | 0.23 | 0.15 | -0.09 |

***Bold** font indicates statistically significant p-values after the correction for multiple comparisons, MDD – major depressive disorder, HC – healthy controls, F – frontal, C – central, P – parietal, O – occipital, T – temporal electrodes, REM – rapid eye movement sleep, N – non–rapid eye movement sleep.*

*Unmedicated vs 7d medicated states*

Both unmedicated and 7d medicated states showed comparable variability of low-band slopes (Fig. S1.1, Table S1.2).

#### Means of high-band slopes

*Unmedicated patients vs controls*

Unmedicated patients and controls showed comparable high-band slopes (Fig. S1.2, Table S1.1).

*7d medicated patients vs controls*

The three-way ANCOVA revealed area-group (F=3.1, p=0.03) and area-stage (F=5.1, p<0.001) interactions. The two-way ANCOVA revealed a main effect with area-group interactions for the N1 (F=7.1, p=0.010), N2 (F=13.3, p<0.001) and N3 (F=10.9, p=0.001) stages.

The *post hoc* analysis revealed that medicated patients showed flatter slopes during N1, N2, and N3 sleep in all areas with moderate to large effect sizes compared to controls. During N1 and N2 sleep, the difference between the groups was greater in the temporal area, and during N3 sleep – in the central electrodes compared to other areas. The wake and REM-related slopes were similar to those measured in the control group (Fig. S1.2, Table S1.1).

*Unmedicated vs 7d medicated states*

The three-way ANOVA revealed a triple area-state-stage interaction (F=2.3, p=0.045). The two-way ANCOVA revealed a main effect with area-state interactions for the N1 (F=7.6, p=0.008), N2 (F=13.7, p=0.001), N3 (F=13.2, p=0.001), and REM (F=20.3, p<0.001) stages.

The *post hoc* analysis revealed that patients showed flatter slopes when medicated than when unmedicated during N1, N2, N3, and REM sleep in all areas with moderate effect sizes. During the N1, N2, and N3 stages, the difference between the states was greater in the temporal area, whereas during REM sleep – in the occipital electrodes compared to other areas. Slopes during the wake epochs were comparable (Fig. S2, Table S1).

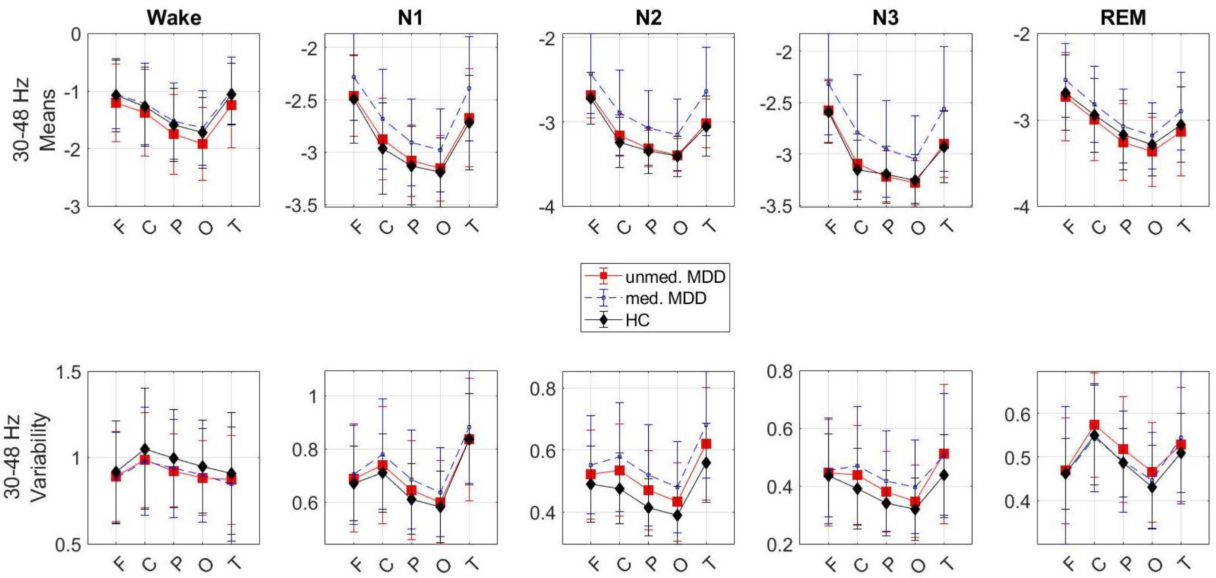


**Figure S1.2. Means and variability of high-band slopes.** Means and intra-individual variability of the slopes of the aperiodic power component in the high (30-48Hz) frequency band over each sleep stage over each area of interest in each study group. **Top:** 7d medicated patients (blue) show flatter slopes compared to the own unmedicated state (red) and controls (black) during all sleep stages in all areas. **Bottom:** Intra-individual variability of slopes was comparable. MDD – 38 major depressive disorder patients, unmed. – unmedicated, med. - 7d medicated, HC – 38 healthy controls, F – frontal, C – central, P – parietal, O – occipital, T – temporal electrodes.

#### Variability of high-band slopes

*Unmedicated vs controls*

Unmedicated patients and controls showed comparable variability of high-band slopes (Fig. S1.2, Table S1.2).

*7d medicated patients vs controls*

The three-way ANCOVA revealed an area-stage interaction (F=6.4, p<0.001). The two-way ANCOVA revealed a main effect of the group on the variability of slopes without a group-area interaction during the N2 stage (F=10.6, the obtained p=0.002 < the corrected threshold p=0.010). The *post hoc* analysis revealed that medicated patients showed greater variability of slopes during N2 sleep in the central, parietal, occipital, and temporal areas with moderate to large effect sizes compared to controls. Variability of slopes during the wake, N1, N3, and REM epochs was comparable (Fig. S1.2, Table S1.2).

*Unmedicated vs 7d medicated states*

Unmedicated and medicated states showed comparable variability of the high-band slopes (Fig. S1.2, Table S1.2).
