## Supplementary Material S2 for "Increased aperiodic neural activity during sleep in major depressive disorder"

Martin Dresler [1,2]

[1]Radboud University, Donders Institute for Brain, Cognition and Behavior, Nijmegen, Netherlands,
[2]Radboud University Medical Centre, Department of Cognitive Neuroscience, Nijmegen, Netherlands,
[3]Netherlands Institute for Neuroscience, Department of Sleep and Cognition, Amsterdam, Netherlands,
[4]Max Planck Institute of Psychiatry, Munich, Germany, [5]Klinikum Ingolstadt, Centre of Mental Health, Ingolstadt, Germany

### Supplementary Material S2

#### Aperiodic activity during resting state

Here, we report resting-state EEG to explore whether the reported in the Main text effects are specific to sleep or can be observable during wake as well.

##### Methods

Resting-state EEG was available in a subset of 16 unmedicated patients and 16 age-matched controls. The signal was recorded in the morning (~ 8 a.m.) following the patient's sleep in the lab. The participants were asked to close their eyes and sit quietly for 15 minutes, staying relaxed in a state of mind-wandering (i.e., no goal-oriented mental activity). Data was filtered using a band-pass finite impulse response filter 0.2-48Hz and divided into 1s epochs. Epochs with voltage > 50μV from the mean across all epochs were rejected from further analysis to reduce noise. Aperiodic component was calculated as described in the Methods of the Main Text and analyzed using three-way ANCOVA with the five-level "brain area" and six-level "stage" (rest, wake epochs of sleep, N1, N2, N3, and REM) as within-subject factors, the two-level "study group" (unmedicated patients and controls) as between-subjects factor, and "age" as a covariate. The Benjamini-Hochberg's adjustment was applied to control for multiple comparisons with a false discovery rate set at 0.05. The corrections were done per 6 tests reflecting the number of sleep stages + resting state with the α-level set in the 0.008–0.050 range.

#### Results

##### *Means of the broadband slopes*

The three-way ANCOVA revealed a significant triple group-area-stage interaction ($F=2.3$, $p=0.037$). The two-way ANCOVA revealed a significant group-area interaction ($F=5.2$, $p=0.003$) during N2 sleep. During the resting state, N1, N3, REM, and wake epochs of sleep there were neither interactions nor main effects of the group on the slopes (Fig. S2.1).

##### *Means of the low-band slopes*

The three-way ANCOVA revealed a significant group-stage interaction (F=3.1, p=0.039). The two-way ANCOVA revealed a significant group-area interaction during N3 sleep (F=5.3, p=0.006). During the resting state, there was a significant main effect of the group on the slope (without an interaction), which, however, did not pass the correction for multiple comparisons (F=4.0, the obtained p=0.054 > the corrected threshold p=0.020). During N1, N2, REM, and wake epochs of sleep, there were neither interactions nor main effects of the group on the slopes. The *post hoc* analysis revealed that unmedicated patients showed flatter slopes during N3 sleep in the frontal areas with a moderate effect size (d=0.7), and steeper slopes during the resting state in the frontal (d=-0.7), central (d=-0.7), and temporal (d=-0.8) electrodes with moderate to large effect sizes compared to controls (Fig. S2.1).

*Means of the high-band slopes*

During rest, unmedicated patients and controls showed comparable high-band slopes.

*Variability of slopes*

During rest, unmedicated patients and controls showed comparable variability of slopes in the low, high, and broadband.

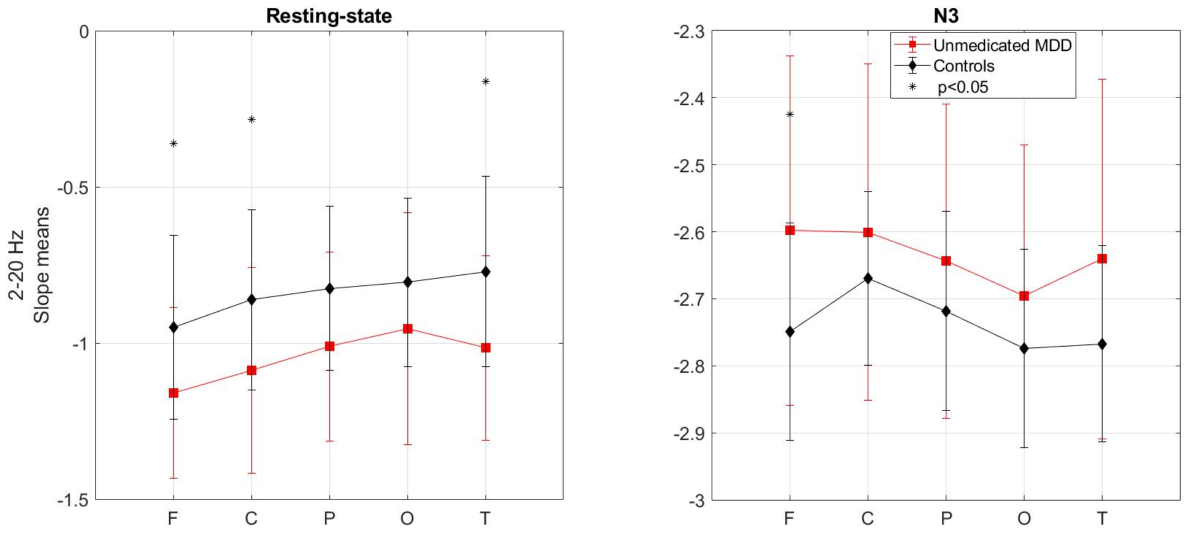


**Figure S2.1. Low-band aperiodic slopes during rest and N3 sleep.** Mean slopes of the low-frequency band aperiodic power (2-20 Hz) were measured during the resting state EEG in the morning in a subset of 16 unmedicated MDD patients (red) and 16 controls (black). Whereas during N3 **(right)**, unmedicated patients (from this subset as well as from the full sample shown in Fig. S1.1) show flatter slopes (more positive values) than controls at the frontal electrodes, the direction of the difference is opposite for the resting state **(left)**, where the patients show steeper slopes (more negative values) than controls at the frontal, central, and temporal electrodes. MDD – major depressive disorder patients, F – frontal, C – central, P – parietal, O – occipital, T – temporal electrodes, asterisks mark statistically significant p-values.
