## Supplementary Material S3 for "Increased aperiodic neural activity during sleep in major depressive disorder"

Martin Dresler [1,2]

[1]Radboud University, Donders Institute for Brain, Cognition and Behavior, Nijmegen, Netherlands,
[2]Radboud University Medical Centre, Department of Cognitive Neuroscience, Nijmegen, Netherlands,
[3]Netherlands Institute for Neuroscience, Department of Sleep and Cognition, Amsterdam, Netherlands,
[4]Max Planck Institute of Psychiatry, Munich, Germany, [5]Klinikum Ingolstadt, Centre of Mental Health, Ingolstadt, Germany

### Supplementary Material S3

#### Aperiodic activity and sleep quality

Based on the literature reporting steeper slopes during deeper sleep stages (1, 2), we hypothesized that aperiodic slopes might reflect subjective sleep quality such that steeper slopes are associated with better sleep quality. We assessed correlations between aperiodic slopes and Pittsburgh Sleep Quality Index (PSQI). PSQI assesses sleep quality and disturbances over a one-month time interval, and, therefore, is not a questionnaire of choice to correlate with a highly dynamic aperiodic activity. Unfortunately, other sleep questionnaires were not available in this study due to its retrospective nature. Nevertheless

#### Methods

A subset of 31 unmedicated patients filled in the PSQI, a self-rated questionnaire where a higher score reflects a worse subjective sleep quality. Associations between the aperiodic slopes and PSQI scores were assessed with Spearman's correlation coefficients. Spearman's correlation was chosen based on visual inspection that revealed the presence of several outliers.

#### Results

For none of the full-length sleep stages (i.e., non-differentiated by early vs late epochs), the slopes did not correlate with the sleep quality scores of the unmedicated patients. After the stratification of N2 by early and late epochs, the slopes showed a trend for correlations with sleep quality for both late and early epochs such that flatter slopes were associated with better sleep quality (lower scores) of unmedicated patients. However, these effects did not reach statistical significance (Fig. S3.1). Unfortunately, we could not stratify other sleep stages by early vs late epochs as we had not enough (only 0–7 epochs) evening or morning epochs for the comparison.

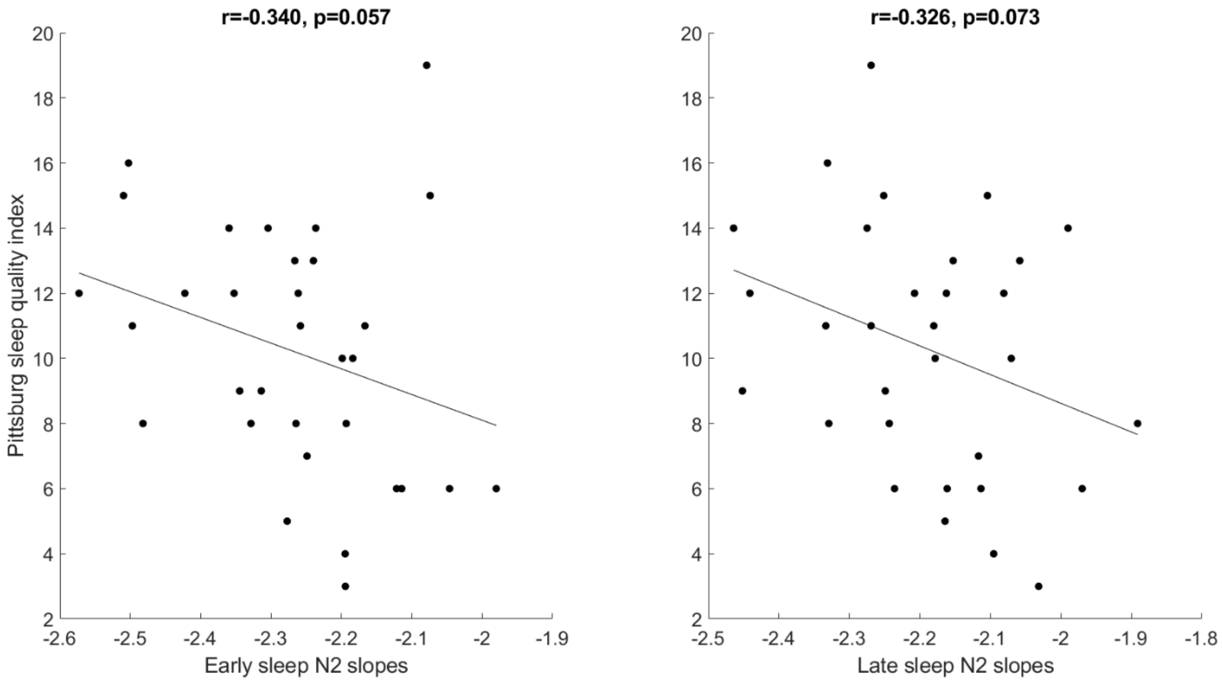


**Figure S3.1. Correlations between early and late N2's aperiodic slopes and sleep quality.** The slopes of the frontal broadband (0.2–48 Hz) aperiodic power component averaged over early **(left)** or late **(right)** non-REM 2 sleep epochs *vs* the Pittsburgh Sleep Quality Index in unmedicated patients (raw values). The flatter slopes of both early and late non-REM sleep 2 are associated with better sleep quality of the unmedicated patients (lower Pittsburgh Sleep Quality Index scores). However, these effects do not reach the statistical significance, r – Spearman's correlations coefficients, n = 31.
