## Supplementary Material S4 for "Increased aperiodic neural activity during sleep in major depressive disorder"

Martin Dresler [1,2]

[1]Radboud University, Donders Institute for Brain, Cognition and Behavior, Nijmegen, Netherlands,
[2]Radboud University Medical Centre, Department of Cognitive Neuroscience, Nijmegen, Netherlands,
[3]Netherlands Institute for Neuroscience, Department of Sleep and Cognition, Amsterdam, Netherlands,
[4]Max Planck Institute of Psychiatry, Munich, Germany, [5]Klinikum Ingolstadt, Centre of Mental Health, Ingolstadt, Germany

### Supplementary Material 4

#### Replication study

To confirm the results reported in the Main study, here, we replicated the analysis reported in the Main Text using two independently collected datasets of short and long-term medicated MDD patients and healthy controls.

##### Methods

These datasets are described in our previous report, where Replication Dataset 1 is referred to as "Dataset C" (thirty MDD patients at 7d and 28d of medication treatment vs thirty healthy controls) and Replication Dataset 2 is referred to as "Dataset A" (forty long-termed medicated MDD patients vs forty healthy controls) (1).

Similar to the main study, here, we calculated the means and intra-individual variability of the aperiodic slopes and compared them between the groups. In addition, we stratified the 7d medicated patients from Replication Dataset 1 into two subgroups by 1) the medication class: SNRIs (n = 12) and TCAs (n = 10); 2) REM-suppressive (n = 21) vs non-suppressive (n = 9) antidepressant type. In Replication Dataset 1, the number of patients who took SSRIs (n = 2), NDRIs (n = 3), or NaSSA (n = 2) was too small, therefore, these subgroups were not analyzed separately. The effect of medication was not analyzed for Replication Dataset 2 since all patients were long-term medicated and most of them took a combination of different drugs.

#### Results

The demographic, clinical, and sleep characteristics of the participants are reported in our previous paper, where the replication dataset 1 is referred to as "Dataset C" and the replication dataset 2 is referred to as "Dataset A" (1).

##### S4.1. Replication dataset 1

###### Means of the broadband slopes

*7d medicated patients vs controls*

The three-way ANCOVA (with the "area", "sleep stage" and "group" factors) revealed a triple area-stage-group interaction between the factors (F=2.5, p=0.022). After the correction for multiple comparisons (five two-way ANCOVAs with the "area" and "group" factors for each sleep stage separately), there was a significant main effect of the group with a group-area interaction on the aperiodic slopes measured during N2 ($F=17.5$, $p<0.001$) and N3 ($F=35.6$, $p<0.001$) sleep, and without a group-area interaction – during N1 ($F=11.8$, $p=0.001$) and REM ($F=12.2$, $p=0.001$) sleep. *Post hoc* analysis revealed that 7d medicated patients showed flatter slopes during the N1, N2, N3, and REM stages in all areas with moderate to very large effect sizes compared to controls. During N2 sleep, the difference between the groups was greater in the frontal and temporal electrodes compared to other areas, whereas during N3 sleep, the difference between the groups was greater in the central electrodes compared to other areas. Slopes of the wake epochs were comparable (Fig. S4.1, Table S4.1).

*28d medicated patients vs controls*

The three-way ANCOVA revealed a significant main effect of the group ($F=11.8$, $p<0.001$) with an area-stage interaction ($F=11.3$, $p<0.001$). The two-way ANCOVA revealed a main effect of the group with a group-area interaction on the slopes measured during N2 ($F=10.2$, $p=0.002$) and N3 ($F=30.1$, $p<0.001$) sleep, and without a group-area interaction – during N1 ($F=7.8$, $p=0.007$) and REM ($F=12.6$, $p=0.001$) sleep. The *post hoc* analysis revealed that 28d medicated patients showed flatter slopes during the N1, N2, N3, and REM stages in all areas with moderate to very large effect sizes compared to controls. During N2 sleep, the difference between the groups was greater in the temporal electrodes compared to other areas, whereas during N3 sleep, the difference between the groups was greater in the central and temporal electrodes compared to other areas. Slopes of the wake epochs were comparable (Fig. S4.1, Table S4.1).

#### Means of the low-band slopes

*7d medicated patients vs controls*

The three-way ANCOVA revealed a triple area-stage-group interaction between the factors (F=4.5, p=0.001). The two-way ANCOVA revealed a main effect of the group without a group-area interaction on the slopes measured during N3 sleep (F=21.9, p<0.001). The *post hoc* analysis revealed that 7d medicated patients showed flatter slopes during N3 sleep in all areas with large effect sizes compared to controls. Slopes of the wake, N1, N2, and REM epochs were comparable (Fig. S4.1, Table S4.1).

*28d medicated patients vs controls*

The three-way ANCOVA revealed a significant triple area-stage-group interaction between the factors (F=4.0, p=0.002). The two-way ANCOVA revealed a main effect of the group without a group-area interaction on the slopes measured during N3 sleep (F=26.2, p<0.001). The *post hoc* analysis revealed that 28d medicated patients showed flatter slopes during N3 sleep in all areas with large effect sizes compared to the controls. Slopes of the wake, N1, N2, and REM epochs were comparable (Fig. S4.1, Table S4.1).

#### Means of the high-band slopes

*7d medicated patients vs controls*

The three-way ANCOVA revealed a significant main effect of the group (F=18.7, p<0.001) with an area-stage interaction (F=8.1, p<0.001). The two-way ANCOVA revealed a significant main effect of the group without group-area interactions on the slopes measured during N1, N2, N3 (all F-values >18.7, all p-values<0.001), and REM (F=10.9, p=0.002) sleep. The *post hoc* analysis revealed that 7d medicated patients showed flatter slopes during N1, N2, N3, and REM sleep in all areas with moderate to very large effect sizes compared to controls. Slopes of the wake epochs were comparable (Fig. S4.1, Table S4.1).

*28d medicated patients vs controls*

The three-way ANCOVA revealed a main effect of the group (F=15.0, p<0.001) with an area-stage interaction (F=7.5, p<0.001). The two-way ANCOVA revealed a main effect of the group without group-area interactions on the slopes measured during the N1, N2, REM (F-values>16.3, p-values<0.001), N3 (F=8.2, p=0.006), and wake (F=4.7, p=0.034) epochs. *Post hoc* analysis revealed that 28d medicated patients showed flatter slopes during all stages in all areas with moderate to very large effect sizes as compared to controls (Fig. S4.1, Table S4.1).

#### Table S4.1: Replication dataset 1. Slope means

| Mean | | 7d medicated MDD | | | | | 28d medicated MDD | | | | | HC | | | | |
|---|---|---|---|---|---|---|---|---|---|---|---|---|---|---|---|---|
| Broadband | Area/ Stage | F | C | P | O | T | F | C | P | O | T | F | C | P | O | T |
| | Wake | -1.41 | -1.29 | -1.63 | -1.71 | -1.33 | -1.39 | -1.28 | -1.65 | -1.74 | -1.33 | -1.50 | -1.38 | -1.70 | -1.76 | -1.42 |
| | N1 | -1.97 | -1.86 | 2.00 | -2.12 | -1.93 | -2.00 | -1.90 | -2.03 | -2.15 | -1.97 | -2.13 | -2.02 | -2.13 | -2.24 | -2.12 |
| | N2 | -2.44 | -2.33 | -2.42 | -2.50 | -2.37 | -2.49 | -2.39 | -2.46 | -2.56 | -2.41 | -2.62 | -2.49 | -2.52 | -2.65 | -2.59 |
| | N3 | -2.76 | -2.64 | -2.70 | -2.78 | -2.68 | -2.78 | -2.67 | -2.65 | -2.81 | -2.69 | -3.00 | -2.87 | -2.87 | -3.00 | -2.98 |
| | REM | -2.01 | -1.89 | -1.98 | -2.09 | -2.05 | -2.01 | -1.89 | -1.97 | -2.09 | -2.02 | -2.10 | -2.01 | -2.13 | -2.24 | -2.17 |
| Low band | Area/ Stage | F | C | P | O | T | F | C | P | O | T | F | C | P | O | T |
| | Wake | -0.95 | -0.77 | -0.99 | -1.10 | -0.87 | -1.01 | -0.85 | -1.08 | -1.19 | -0.94 | -0.99 | -0.79 | -0.97 | -1.00 | -0.86 |
| | N1 | -1.41 | -1.25 | -1.37 | -1.54 | -1.43 | -1.44 | -1.29 | -1.40 | -1.58 | -1.45 | -1.52 | -1.35 | -1.46 | -1.62 | -1.52 |
| | N2 | -1.92 | -1.79 | -1.89 | -2.03 | -1.93 | -1.96 | -1.84 | -1.92 | -2.06 | -1.96 | -2.05 | -1.89 | -1.95 | -2.12 | -2.04 |
| | N3 | -2.37 | -2.24 | -2.32 | -2.41 | -2.37 | -2.34 | -2.23 | -2.22 | -2.39 | -2.33 | -2.62 | -2.47 | -2.50 | -2.62 | -2.61 |
| | REM | -1.61 | -1.41 | -1.46 | -1.56 | -1.56 | -1.59 | -1.39 | -1.44 | -1.55 | -1.53 | -1.62 | -1.44 | -1.56 | -1.64 | -1.60 |
| High band | Area/ Stage | F | C | P | O | T | F | C | P | O | T | F | C | P | O | T |
| | Wake | -2.76 | -2.82 | -3.28 | -3.30 | -2.61 | -2.68 | -2.68 | -3.15 | -3.27 | -2.58 | -3.01 | -3.08 | -3.59 | -3.69 | -2.90 |
| | N1 | -3.91 | -3.90 | -4.08 | -4.03 | -3.68 | -3.91 | -3.91 | -4.07 | -4.11 | -3.76 | -4.40 | -4.38 | -4.50 | -4.57 | -4.25 |
| | N2 | -3.97 | -3.91 | -4.05 | -4.05 | -3.78 | -3.94 | -3.92 | -4.01 | -4.07 | -3.78 | -4.43 | -4.33 | -4.38 | -4.55 | -4.32 |
| | N3 | -3.90 | -3.78 | -3.90 | -3.98 | -3.71 | -3.91 | -3.83 | -3.60 | -3.98 | -3.80 | -4.21 | -4.09 | -4.09 | -4.31 | -4.11 |
| | REM | -3.90 | -3.98 | -4.10 | -4.28 | -4.09 | -3.87 | -3.96 | -4.04 | -4.23 | -4.04 | -4.32 | -4.37 | -4.47 | -4.70 | -4.54 |
| **Effect size** | | **7d medicated MDD-HC** | | | | | **28d medicated MDD-HC** | | | | | **7d medicated-28d medicated MDD** | | | | |
| Broadband | Area/ Stage | F | C | P | O | T | F | C | P | O | T | F | C | P | O | T |
| | W | 0.32 | 0.24 | 0.24 | 0.22 | 0.30 | 0.32 | 0.27 | 0.16 | 0.07 | 0.27 | -0.05 | -0.04 | 0.10 | 0.19 | 0.01 |
| | N1 | **0.90** | **0.74** | **0.70** | **0.81** | **0.90** | **0.74** | **0.65** | **0.55** | **0.55** | **0.76** | 0.19 | 0.25 | 0.22 | **0.38** | 0.27 |

| | | | | | | | | | | | | | | | | |
|---|---|---|---|---|---|---|---|---|---|---|---|---|---|---|---|---|
| | N2 | **1.05** | **0.97** | **0.75** | **0.97** | **1.07** | **0.85** | **0.71** | 0.42 | **0.57** | **0.97** | 0.34 | **0.42** | 0.29 | **0.39** | **0.38** |
| | N3 | **1.24** | **1.42** | **1.12** | **1.31** | **1.32** | **1.15** | **1.28** | **0.94** | **1.00** | **1.30** | 0.14 | 0.21 | 0.14 | 0.26 | 0.05 |
| | REM | **0.75** | **0.74** | **0.95** | **0.92** | **0.78** | **0.75** | **0.74** | **0.94** | **0.88** | **0.91** | -0.03 | -0.01 | -0.04 | 0.02 | -0.26 |
| Low band | Area/ Stage | F | C | P | O | T | F | C | P | O | T | F | C | P | O | T |
| | Wake | 0.13 | 0.07 | -0.06 | -0.33 | -0.02 | -0.01 | -0.08 | -0.31 | **-0.63** | -0.16 | 0.18 | 0.19 | 0.43 | **0.36** | 0.18 |
| | N1 | 0.48 | 0.41 | 0.35 | 0.32 | 0.41 | 0.39 | 0.28 | 0.25 | 0.20 | 0.35 | 0.17 | 0.23 | 0.16 | 0.21 | 0.12 |
| | N2 | **0.65** | 0.49 | 0.30 | 0.43 | **0.51** | 0.44 | 0.25 | 0.15 | 0.29 | 0.41 | **0.36** | **0.42** | 0.23 | 0.24 | 0.21 |
| | N3 | **1.15** | **1.08** | **0.88** | **1.00** | **1.14** | **1.19** | **1.15** | **1.01** | **1.08** | **1.34** | -0.12 | -0.11 | -0.12 | -0.08 | -0.30 |
| | REM | 0.06 | 0.15 | 0.42 | 0.34 | 0.15 | 0.14 | 0.23 | 0.49 | 0.38 | 0.29 | -0.13 | -0.19 | -0.14 | -0.10 | -0.36 |
| High band | Area/ Stage | F | C | P | O | T | F | C | P | O | T | F | C | P | O | T |
| | Wake | 0.33 | 0.30 | 0.41 | **0.55** | 0.38 | 0.46 | 0.50 | **0.60** | **0.63** | 0.48 | -0.21 | -0.26 | -0.27 | -0.16 | -0.16 |
| | N1 | **0.91** | **0.87** | **1.00** | **1.23** | **0.99** | **0.94** | **0.95** | **1.00** | **1.05** | **0.90** | -0.13 | -0.07 | -0.12 | 0.08 | 0.04 |
| | N2 | **1.10** | **1.07** | **1.03** | **1.23** | **1.11** | **1.11** | **1.05** | **1.01** | **1.12** | **1.10** | -0.20 | -0.06 | -0.19 | -0.02 | -0.16 |
| | N3 | **0.69** | **0.79** | **0.62** | **0.84** | **0.76** | **0.66** | **0.65** | **0.56** | **0.77** | **0.67** | -0.07 | 0.04 | -0.07 | -0.09 | 0.02 |
| | REM | **1.04** | **0.97** | **1.07** | **1.35** | **1.20** | **1.13** | **1.04** | **1.14** | **1.30** | **1.13** | -0.21 | -0.19 | -0.25 | -0.26 | -0.20 |

***Bold** font indicates statistically significant p-values after the correction for multiple comparisons, MDD – major depressive disorder, HC – healthy controls, W – wake, F – frontal, C – central, P – parietal, O – occipital, T – temporal electrodes, REM – rapid eye movement sleep, N – non–rapid eye movement sleep.*

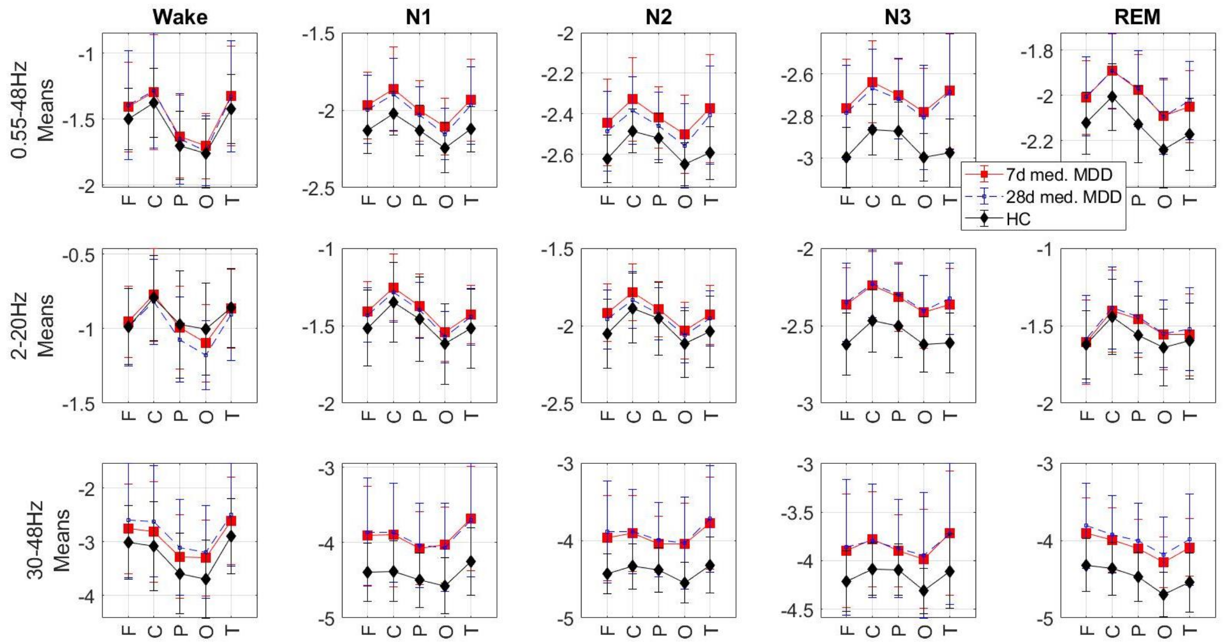


**Figure S4.1. Replication dataset 1: means of slopes.** The slopes of the aperiodic power component in the 0.55-48 Hz **(top)**, 2-20 Hz **(middle)**, and 30-48 Hz **(bottom)** were averaged over each sleep stage over each area of interest. Patients at 7d (red) and 28d (blue) of medication treatment show flatter slopes (more positive values) of the high-band (top) and broadband (top) power than controls (black) in all areas during all sleep stages. For the low band (middle), the difference was prominent only during N3 sleep. MDD – 30 major depressive disorder, HC – 30 healthy controls, F – frontal, C – central, P – parietal, O – occipital, T – temporal electrodes.

#### Medication effect

To replicate the finding on the specific effect of the antidepressant class observed in the Main Text, we stratified the 7d medicated patients from Replication Dataset 1 by their medication class. In contrast to the Main dataset, we found that aperiodic activity was comparable in the patients who took REM-suppressive and REM non-suppressive antidepressants as well as in patients who took SNRIs and non-SNRIs or other than SNRIs REM-suppressive antidepressants. The most probable explanation of this discrepancy is that in the main study, patients took different REM suppressive agents than in the replication study, and it is well-known that different agents even from the same class can show opposite modes of action on sleep (2). Moreover, in clinical settings, response to antidepressants is highly variable (2). Other reasons include the older age and higher number of depression episodes (that might correspond to the disorder progression) of the patients from the replication dataset. Likewise, the replication analysis had low statistical power and unbalanced groups (21 vs 9 patients in the replication dataset compared to 21 vs 17 patients in the main study).

At the same time, in the Replication Dataset 1, the patients who took TCA showed flatter aperiodic slopes during REM sleep in the central, parietal, occipital, and temporal areas with moderate to large effect sizes compared to their pooled controls (all p-values ranging from 0.011 to 0.025, all Cohen's d-values ranging from 0.71 to 0.97). However, this finding did not pass the correction for multiple comparisons (25 non-parametric tests for 5 stages by 5 areas separately, Table S4.2).

**Table S4.2: Effect of antidepressants on aperiodic activity**

| **Main study** | | | **Replication study 1** | | |
|---|---|---|---|---|---|
| **Subgroups** | | **Slope difference** | **Subgroups** | | **Slope difference** |
| 13 SSRI | 25 non-SSRI | n.s. | 2 SSRI | 28 non-SSRI | NA |
| 8 TCA | 30 non-TCA | n.s. | 10 TCA | 20 non-TCA | ↑ REM<br>C, P, O, T |
| 6 NDRI | 32 non-NDRI | n.s. | 3 NDRI | 27 non-NDRI | NA |
| 6 SNRI | 32 non-SNRI | ↑ N1, **N2, N3, REM**<br>F, C, P, O, T | 12 SNRI | 18 non-SNRI | n.s. |
| 21 REM suppressive | 17 REM non-suppressive | ↑ **N1, N2, N3, REM**<br>F, C, P, O, T | 21 REM suppressive | 9 REM non-suppressive | n.s. |
| 6 SNRI (REM suppressive) | 15 REM suppressive non-SNRI | ↑ REM<br>F, C, P, O, T | 12 SNRI | 18 REM non-suppressive | n.s. |
| 6 SNRI (REM suppressive) | 13 SSRI (REM suppressive) | ↑ REM<br>F, C, P, O, T | 12 SNRI | 2 SSRI | NA |

***Bold** font indicates the finding that remained statistically significant after the correction for multiple comparisons, F – frontal, C – central, P – parietal, O – occipital, T – temporal electrodes, n.s. – non-significant, REM – rapid eye movement sleep, N – non–rapid eye movement sleep, NaSSA – noradrenergic and specific serotonergic antidepressants, NDRI – norepinephrine-dopamine reuptake inhibitor, SNRI – serotonin-norepinephrine reuptake inhibitors, SSRI – selective serotonin reuptake inhibitors, TCA – tricyclic antidepressants*

#### Variability of the broadband slopes

*7d medicated patients vs controls*

The three-way ANCOVA revealed an area-stage interaction (F=2.9, p=0.01). The two-way ANCOVA revealed a main effect of the group (without a group-area interaction) on the variability of slopes during N2 sleep (F=9.1, the obtained p=0.004 < the corrected threshold p=0.010), and a marginally significant main effect during N3 sleep (F=5.3, the obtained p=0.025 > the corrected threshold p=0.020). The *post hoc* analysis revealed that 7d medicated patients showed greater variability of slopes during N2 sleep in all areas and during N3 sleep – in the frontal, central, and occipital areas with moderate effect sizes compared to controls. Variability of slopes of the wake, N1, and REM epochs was comparable (Fig. S4.2, Table S4.3).

*28d medicated patients vs controls*

The three-way ANCOVA (with the "area", "stage" and "group" factors) revealed a significant triple area-stage-group interaction (F=2.5, p=0.017). The two-way ANCOVA revealed a main effect of the group (without a group-area interaction) on the variability of slopes during N2 (F=9.2, the obtained p=0.004 < the corrected threshold p=0.010) and N3 (F=5.9, p=0.019 < the corrected threshold p=0.020) sleep, and a marginally significant main effect during REM sleep (F=4.9, the obtained p=0.031 > the corrected threshold p=0.030). The *post hoc* analysis revealed that 28d medicated patients showed greater variability of slopes during N2, N3, and REM sleep in all areas with moderate to large effect sizes compared to controls. Variability of slopes of the wake and N1 epochs was comparable (Fig. S4.2, Table S4.3).

#### Variability of the low-band slopes

There were no differences between the groups in the variability of the low-frequency band slopes (Fig. S4.2, Table S4.3).

#### Variability of the high-band slopes

*7d medicated patients vs controls*

The three-way ANCOVA revealed the stage-group ($F=3.9$, $p=0.026$) and area-stage ($F=2.7$, $p=0.018$) interactions. The two-way ANCOVA revealed a main effect of the group (without a group-area interaction) on the variability of slopes during N2 ($F=10.5$, the obtained $p=0.002$ < the corrected threshold $p=0.010$), N3 ($F=6.3$, the obtained $p=0.015$ < the corrected threshold $p=0.020$), and N1 ($F=5.2$, the obtained $p=0.026$ < the corrected threshold $p=0.030$) stages. The *post hoc* analysis revealed that 7d medicated patients showed greater variability of slopes during N2 sleep in all areas, during N3 sleep – in the frontal and central areas, and during N1 sleep – in the frontal and occipital areas with moderate effect sizes compared to controls. Variability of slopes of the wake and REM epochs was comparable (Fig. S4.2, Table S4.3).

*28d medicated patients vs controls*

The three-way ANCOVA revealed the main effects of the area ($F=6.4$, $p<0.001$), stage ($F=16.1$, $p<0.001$), and group ($F=7.9$, $p=0.007$) without interactions. The two-way ANCOVA revealed a main effect of the group (without a group-area interaction) on the variability of slopes during N2 ($F=10.7$, the obtained $p=0.002$ < the corrected threshold $p=0.010$), N3 ($F=8.6$, the obtained p=0.005 < the corrected threshold p=0.020), and N1 sleep (F=5.0, the obtained p=0.029 < the corrected threshold p=0.030). The *post hoc* analysis revealed that 28d medicated patients showed greater variability of slopes during N2 and N3 sleep in all areas, and during N1 sleep – in the frontal and central areas with moderate to large effect sizes compared to controls. Variability of slopes of the wake and REM epochs was comparable (Fig. S4.2, Table S4.3).

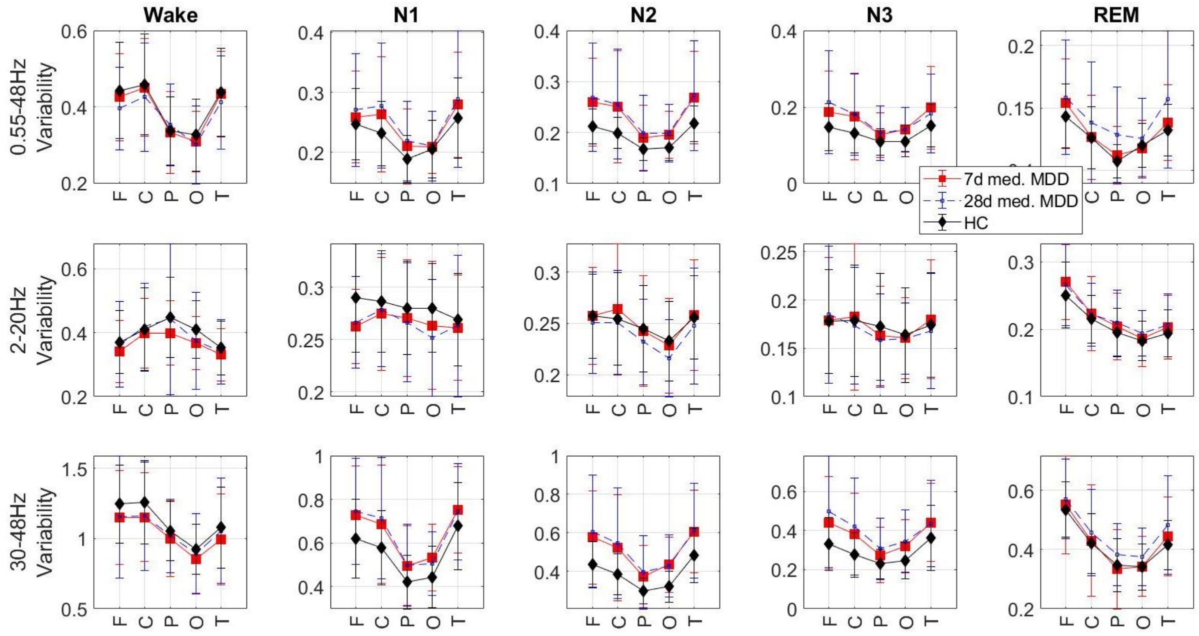


**Figure S4.2. Replication dataset 1: variability of slopes.** Intra-individual variability of the slopes of the aperiodic power component in the 0.55-48 Hz **(top)**, 2-20 Hz **(middle)**, and 30-48 Hz **(bottom)** bands was averaged over each sleep stage over each area of interest. Patients at 7d (red) and 28d (blue) of medication treatment show greater intra-individual variability of the slopes of the high (bottom) and broadband (top) but not low-band (middle) power compared to controls (black) during N2 and N3 sleep. MDD – 30 patients with major depressive disorder, HC – 30 healthy controls, F – frontal, C – central, P – parietal, O – occipital, T – temporal electrodes.

**Table S4.3: Replication dataset 1. Intra-individual variability of slopes**

| Std | | 7d medicated MDD | | | | | 28d medicated MDD | | | | | HC | | | | |
|---|---|---|---|---|---|---|---|---|---|---|---|---|---|---|---|---|
| Broadband | Area/ Stage | F | C | P | O | T | F | C | P | O | T | F | C | P | O | T |
| | Wake | 0.43 | 0.45 | 0.33 | 0.31 | 0.43 | 0.40 | 0.43 | 0.35 | 0.30 | 0.41 | 0.44 | 0.46 | 0.34 | 0.33 | 0.44 |
| | N1 | 0.26 | 0.26 | 0.21 | 0.21 | 0.28 | 0.27 | 0.28 | 0.22 | 0.21 | 0.29 | 0.25 | 0.23 | 0.19 | 0.21 | 0.26 |
| | N2 | 0.26 | 0.25 | 0.19 | 0.20 | 0.27 | 0.27 | 0.26 | 0.20 | 0.20 | 0.27 | 0.21 | 0.20 | 0.17 | 0.17 | 0.22 |
| | N3 | 0.19 | 0.17 | 0.13 | 0.14 | 0.20 | 0.21 | 0.18 | 0.16 | 0.14 | 0.18 | 0.15 | 0.13 | 0.11 | 0.11 | 0.15 |
| | REM | 0.15 | 0.13 | 0.11 | 0.12 | 0.14 | 0.16 | 0.14 | 0.13 | 0.13 | 0.16 | 0.14 | 0.13 | 0.11 | 0.12 | 0.13 |
| Low band | Area/ Stage | F | C | P | O | T | F | C | P | O | T | F | C | P | O | T |
| | Wake | 0.34 | 0.40 | 0.40 | 0.37 | 0.33 | 0.36 | 0.42 | 0.44 | 0.37 | 0.34 | 0.37 | 0.41 | 0.45 | 0.41 | 0.35 |
| | N1 | 0.26 | 0.28 | 0.27 | 0.26 | 0.26 | 0.27 | 0.28 | 0.27 | 0.25 | 0.26 | 0.29 | 0.29 | 0.28 | 0.28 | 0.27 |
| | N2 | 0.26 | 0.26 | 0.24 | 0.23 | 0.26 | 0.25 | 0.25 | 0.23 | 0.22 | 0.25 | 0.26 | 0.25 | 0.25 | 0.23 | 0.26 |
| | N3 | 0.18 | 0.18 | 0.16 | 0.16 | 0.18 | 0.19 | 0.17 | 0.13 | 0.16 | 0.17 | 0.18 | 0.18 | 0.17 | 0.16 | 0.17 |
| | REM | 0.27 | 0.22 | 0.20 | 0.19 | 0.20 | 0.27 | 0.22 | 0.21 | 0.19 | 0.21 | 0.25 | 0.22 | 0.20 | 0.18 | 0.19 |
| High band | Area/ Stage | F | C | P | O | T | F | C | P | O | T | F | C | P | O | T |
| | Wake | 1.15 | 1.15 | 1.00 | 0.86 | 0.99 | 1.15 | 1.16 | 1.02 | 0.89 | 1.05 | 1.25 | 1.26 | 1.05 | 0.92 | 1.08 |
| | N1 | 0.73 | 0.69 | 0.50 | 0.53 | 0.75 | 0.75 | 0.71 | 0.50 | 0.51 | 0.74 | 0.62 | 0.58 | 0.42 | 0.44 | 0.68 |
| | N2 | 0.58 | 0.52 | 0.37 | 0.44 | 0.61 | 0.61 | 0.55 | 0.40 | 0.43 | 0.61 | 0.44 | 0.39 | 0.30 | 0.32 | 0.48 |
| | N3 | 0.44 | 0.38 | 0.28 | 0.32 | 0.44 | 0.50 | 0.42 | 0.35 | 0.35 | 0.44 | 0.33 | 0.28 | 0.23 | 0.25 | 0.36 |
| | REM | 0.55 | 0.43 | 0.33 | 0.34 | 0.44 | 0.57 | 0.46 | 0.38 | 0.38 | 0.48 | 0.54 | 0.42 | 0.35 | 0.34 | 0.42 |
| **Effect size** | | **7d medicated MDD-HC** | | | | | **28d medicated MDD-HC** | | | | | **7d medicated-28d medicated MDD** | | | | |
| Broadband | Area/ Stage | F | C | P | O | T | F | C | P | O | T | F | C | P | O | T |
| | Wake | -0.14 | -0.06 | -0.05 | -0.19 | -0.04 | -0.40 | -0.24 | 0.17 | -0.27 | -0.22 | 0.27 | 0.21 | -0.19 | 0.10 | 0.18 |
| | N1 | 0.18 | 0.42 | 0.41 | 0.08 | 0.29 | 0.31 | 0.57 | 0.55 | 0.09 | 0.34 | -0.23 | -0.20 | -0.17 | -0.01 | -0.12 |

| | | | | | | | | | | | | | | | | |
|---|---|---|---|---|---|---|---|---|---|---|---|---|---|---|---|---|
| | N2 | 0.74 | 0.66 | 0.47 | 0.71 | 0.76 | 0.74 | 0.73 | 0.58 | 0.68 | 0.70 | -0.16 | -0.06 | -0.15 | -0.08 | -0.06 |
| | N3 | 0.45 | 0.51 | 0.38 | 0.61 | 0.56 | 0.63 | 0.61 | 0.41 | 0.62 | 0.39 | -0.33 | -0.08 | -0.07 | 0.00 | 0.17 |
| | REM | 0.34 | **0.01** | **0.28** | -0.12 | 0.24 | 0.42 | 0.32 | 0.76 | 0.23 | 0.62 | **-0.09** | -0.25 | -0.43 | -0.24 | -0.37 |
| Low band | Area/ Stage | F | C | P | O | T | F | C | P | O | T | F | C | P | O | T |
| | Wake | -0.30 | -0.10 | -0.43 | -0.49 | -0.27 | -0.06 | 0.07 | -0.03 | -0.29 | -0.13 | -0.17 | -0.15 | -0.20 | -0.04 | -0.10 |
| | N1 | -0.61 | -0.23 | -0.20 | -0.32 | -0.17 | -0.49 | -0.16 | -0.28 | -0.58 | -0.11 | -0.09 | -0.09 | 0.10 | 0.23 | -0.03 |
| | N2 | 0.01 | 0.18 | -0.04 | -0.10 | 0.05 | -0.14 | -0.08 | -0.31 | -0.43 | -0.17 | 0.15 | 0.29 | 0.22 | 0.32 | 0.25 |
| | N3 | 0.02 | 0.06 | -0.18 | -0.07 | 0.10 | 0.11 | -0.09 | -0.27 | -0.09 | -0.11 | -0.11 | 0.14 | 0.08 | 0.02 | 0.22 |
| | REM | 0.37 | 0.18 | 0.22 | 0.10 | 0.23 | 0.29 | 0.16 | 0.34 | 0.34 | 0.29 | 0.06 | 0.04 | -0.15 | -0.23 | -0.08 |
| High band | Area/ Stage | F | C | P | O | T | F | C | P | O | T | F | C | P | O | T |
| | Wake | -0.31 | -0.35 | -0.24 | -0.32 | -0.27 | -0.25 | -0.29 | -0.15 | -0.14 | -0.07 | -0.01 | -0.02 | -0.11 | -0.16 | -0.20 |
| | N1 | 0.53 | 0.49 | 0.49 | 0.62 | 0.38 | 0.59 | 0.60 | 0.50 | 0.44 | 0.32 | -0.09 | -0.13 | -0.02 | 0.21 | 0.05 |
| | N2 | 0.75 | 0.67 | 0.61 | 0.99 | 0.69 | 0.78 | 0.76 | 0.70 | 0.85 | 0.65 | -0.18 | -0.13 | -0.16 | 0.06 | -0.03 |
| | N3 | 0.57 | 0.64 | 0.40 | 0.65 | 0.44 | 0.75 | 0.76 | 0.63 | 0.75 | 0.38 | -0.35 | -0.26 | -0.29 | -0.18 | 0.03 |
| | REM | 0.12 | 0.06 | -0.12 | 0.01 | 0.26 | 0.31 | 0.29 | 0.38 | 0.38 | 0.52 | -0.15 | -0.22 | -0.46 | -0.40 | -0.29 |

***Bold** font indicates statistically significant p-values after the correction for multiple comparisons, MDD – major depressive disorder, HC – healthy controls, F – frontal, C – central, P – parietal, O – occipital, T – temporal electrodes, REM – rapid eye movement sleep, N – non–rapid eye movement sleep.*

**S4.2. Replication dataset 2**

**Means of the broadband slope**

After the correction for multiple comparisons (five t-tests for each sleep stage separately), the long-term medicated patients showed flatter aperiodic slopes during N2, N3, and REM sleep (all p-values$<0.003$) in the central electrodes (other channels were unavailable in this dataset) with moderate effect sizes compared to controls. Slopes of the wake epochs were comparable (Fig. S4.3, Table S4.4).

**Means of the low-band slope**

The medicated patients showed flatter aperiodic slopes during N1, N2, N3, and REM sleep (all p-values$<0.003$) in the central electrodes with moderate to large effect sizes compared to controls. Slopes of the wake epochs were comparable (Fig. S4.3, Table S4.4).

**Means of the high-band slope**

The medicated patients showed flatter aperiodic slopes during N2, N3, and REM sleep (all p-values$<0.003$) in the central electrodes with moderate effect sizes compared to controls. Slopes of the wake epochs were comparable (Fig. S4.3, Table S4.4).

#### Table S4.4: Replication dataset 2. Means and variability of slopes

| Groups | | Long-term medicated MDD | HC | MDD-HC | Long-term medicated MDD | HC | MDD-HC |
|---|---|---|---|---|---|---|---|
| Band | Stage | Slope means | | Effect size | Slope variability | | Effect size |
| Broadband | Wake | -1.38 | -1.36 | -0.05 | 0.36 | 0.32 | 0.30 |
| | N1 | -1.74 | -1.84 | **0.45** | 0.31 | 0.34 | -0.25 |
| | N2 | -2.15 | -2.34 | **0.70** | 0.28 | 0.29 | -0.12 |
| | N3 | -2.44 | -2.68 | **0.79** | 0.16 | 0.22 | **-0.48** |
| | REM | -1.95 | -2.11 | **0.69** | 0.18 | 0.18 | -0.01 |
| Low band | Wake | -0.86 | -0.83 | -0.09 | 0.33 | 0.31 | 0.15 |
| | N1 | -1.31 | -1.46 | **0.69** | 0.28 | 0.33 | -0.49 |
| | N2 | -1.78 | -1.98 | **0.71** | 0.27 | 0.27 | -0.07 |
| | N3 | -2.18 | -2.44 | **0.95** | 0.14 | 0.22 | **-0.87** |
| | REM | -1.45 | -1.64 | **0.70** | 0.23 | 0.23 | 0.10 |
| High band | Wake | -2.78 | -2.90 | 0.20 | 0.62 | 0.55 | 0.27 |
| | N1 | -3.07 | -3.26 | 0.43 | 0.58 | 0.58 | -0.01 |
| | N2 | -3.08 | -3.37 | **0.66** | 0.46 | 0.46 | 0.00 |
| | N3 | -3.06 | -3.28 | **0.50** | 0.28 | 0.34 | -0.34 |
| | REM | -3.58 | -3.83 | **0.60** | 0.31 | 0.32 | -0.10 |

***Bold*** *font indicates statistically significant p-values, all values were averaged over the central electrodes, MDD – major depressive disorder, HC – healthy controls, REM – rapid eye movement sleep, N – non–rapid eye movement sleep.*

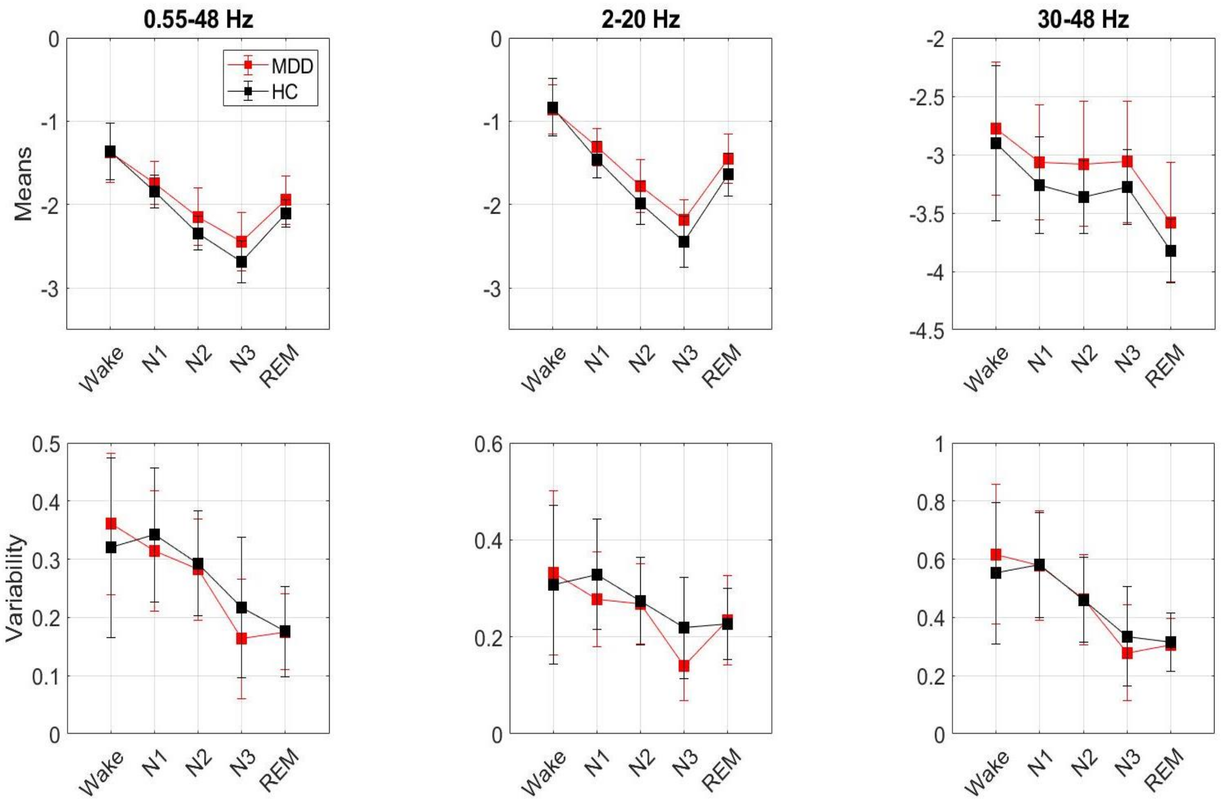


**Figure S4.3. Replication dataset 2: means and variability of slopes.** Means **(top)** and intra-individual variability **(bottom)** of the slopes of the aperiodic power component in the 0.55-48Hz (left), 2-20Hz (middle), and 30-48Hz (right) bands were averaged over each sleep stage over the central electrodes. Long-term medicated patients (red) show flatter slopes (more positive values) during non-REM and REM sleep in all frequency bands (top) and lower variability of slopes during N3 sleep in low-band (bottom middle) and broadband (bottom left) compared to controls (black). MDD – 40 long-term medicated major depressive disorder patients, HC – 40 healthy controls.

#### Variability of the broadband slopes

During N3 sleep, the medicated patients showed lower intra-individual variability of slopes compared to controls in the central electrodes. However, this effect did not pass the correction for multiple comparisons (the obtained p=0.038 > the corrected threshold p=0.010 for 5 separate t-tests). Patients and controls showed comparable variability of slopes of the wake, N1, N2, and REM epochs (Fig. S4.3, Table S4.4).

**Variability of the low-band slopes**

During N3 sleep, the medicated patients showed lower variability of slopes compared to controls in the central electrodes with a large effect size ($p<0.001$, $d=-0.87$). Likewise, patients showed greater variability of slopes during N1 sleep in the central areas. However, this effect did not pass the correction for multiple comparisons (the obtained $p=0.032$ < the corrected threshold $p=0.020$). Patients and controls showed comparable variability of slopes of the wake, N2, and REM epochs (Fig. S4.3, Table S4.4).

**Variability of the high-band slopes**

Both groups showed comparable variability of slopes in the central electrodes (Fig. S4.3, Table S4.4).

#### S4.3. Comparison between the datasets

The comparison between the Main dataset, Replication dataset 1, and Replication Dataset 2 are presented in Fig. S4.4 and Table S4.5. We found that medicated patients from all datasets showed flatter slopes during both REM and non-REM sleep. Medicated patients from the Main study (7d medicated) and Replication dataset 1 (7d and 28d medicated) but not Replication Dataset 2 (long-term medicated) showed greater variability of slopes during non-REM sleep.

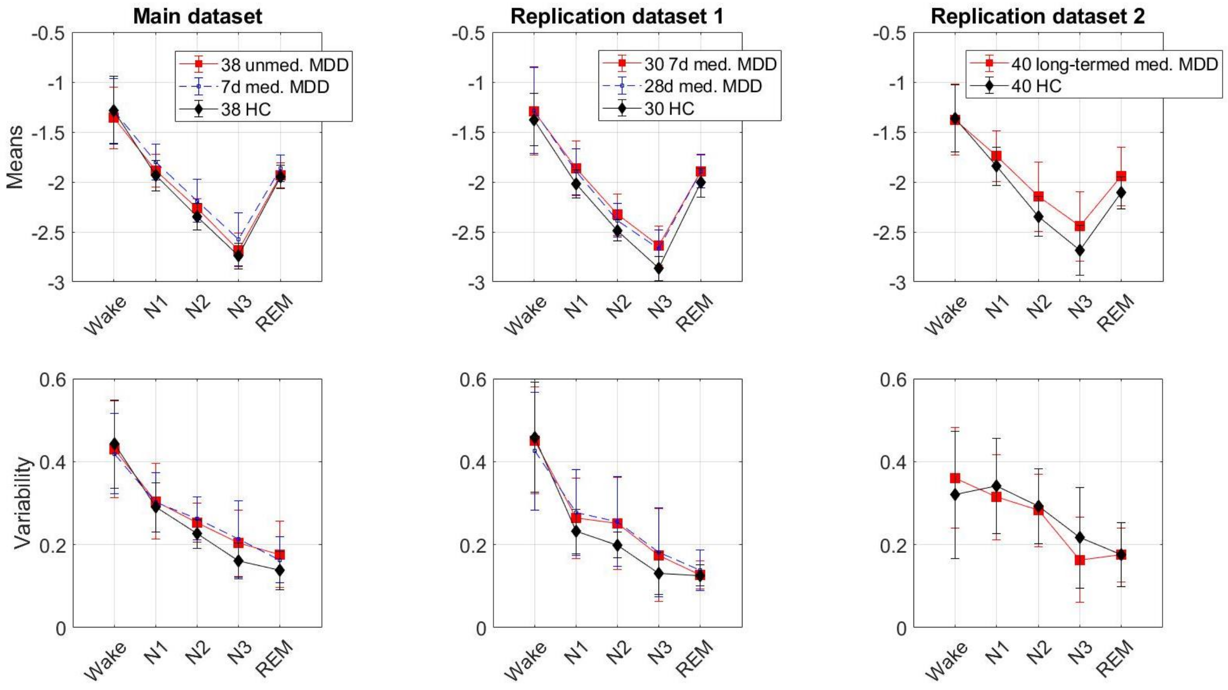


**Figure S4.4. Main and replication datasets comparison.** Means **(top)** and intra-individual variability **(bottom)** of the slopes of the aperiodic broadband power component were averaged over each sleep stage over the central electrodes. **Top:** Unmedicated patients (red, left) show flatter slopes (more positive values) during N2 and N3 sleep compared to controls (black). Medicated patients from all datasets show flatter slopes during both REM and non-REM sleep compared to controls. **Bottom:** Unmedicated patients (red, left) show greater variability of slopes during both non-REM and REM sleep compared to controls (black). Medicated patients from the Main study (7d medicated, blue, left) and Replication dataset 1 (7d and 28d medicated, red and blue, middle) but not Replication Dataset 2 (long-term medicated, red, right) show greater variability of slopes during non-REM sleep (bottom left and central) compared to controls (black). MDD – major depressive disorder patients, HC – healthy controls.

#### Table S4.5: Summary of the *post hoc* findings

| Band | Broadband (0.2-48 Hz) | | Low band (2-20 Hz) | | High band (30-48 Hz) | | Broadband (0.2-48 Hz) | | Low band (2-20 Hz) | | High band (30-48 Hz) | |
|---|---|---|---|---|---|---|---|---|---|---|---|---|
| **Variable** | **Means** | | | | | | **Variability** | | | | | |
| **Main study** | | | | | | | | | | | | |
| **State/ Stage** | **Unmed.** | **7d med.** | **Unmed.** | **7d med.** | **Unmed.** | **7d med.** | **Unmed.** | **7d med.** | **Unmed.** | **7d med.** | **Unmed.** | **7d med.** |
| Wake | n.s. | n.s. | n.s. | n.s. | n.s. | n.s. | n.s. | n.s. | n.s. | n.s. | n.s. | n.s. |
| N1 | n.s. | ↑ F, C, P, O, T | n.s. | ↑ F, C, P, O, T | n.s. | ↑ F, C, P, O, T | n.s. | n.s. | n.s. | n.s. | n.s. | n.s. |
| N2 | ↑ F, C, P, O, T | ↑ F, C, P, O, T | ↑ F, C, P, O, T | ↑ F, C, P, O, T | n.s. | ↑ F, C, P, O, T | ↑ F, C, P, O, T | ↑ F, C, P, O, T | n.s. | n.s. | n.s. | ↑ C, P, O, T |
| N3 | ↑ F, T | ↑ F, C, P, O, T | ↑ F, C, P, O, T | ↑ F, C, P, O, T | n.s. | ↑ F, C, P, O, T | ↑ F, C, P, O, T | ↑ F, C, P, O, T | n.s. | n.s. | n.s. | n.s. |
| REM | n.s. | ↑ F, C, P, O, T | n.s. | ↑ C, P, O, T | n.s. | n.s. | ↑ C, P, O . | ↑ F, C, P, O, T | ↑ F, C, P, O, T | ↑ F, C, P, O, T | n.s. | n.s. |
| **Replication dataset 1** | | | | | | | | | | | | |
| **State/ Stage** | **7d med.** | **28d med.** | **7d med.** | **28d med.** | **7d med.** | **28d med.** | **7d med.** | **28d med.** | **7d med.** | **28d med.** | **7d med.** | **28d med.** |
| Wake | n.s. | n.s. | n.s. | ↑ O | ↑ O | ↑ P, O | n.s. | n.s. | n.s. | n.s. | n.s. | n.s. |
| N1 | ↑ F, C, P, O, T | ↑ F, C, P, O, T | n.s. | n.s. | ↑ F, C, P, O, T | ↑ F, C, P, O, T | n.s. | n.s. | n.s. | n.s. | ↑ F, O | ↑ F, C |
| N2 | ↑ F, C, P, O, T | ↑ F, C, O, T | ↑ F, T | n.s. | ↑ F, C, P, O, T | ↑ F, C, P, O, T | ↑ F, C, P, O, T | ↑ F, C, P, O, T | n.s. | n.s. | ↑ F, C, P, O, T | ↑ F, C, O, T |
| N3 | ↑ F, C, P, O, T | ↑ F, C, P, O, T | ↑ F, C, P, O, T | ↑ F, C, P, O, T | ↑ F, C, P, O, T | ↑ F, C, P, O, T | ↑ F, C, O | ↑ F, C, P, O, T | n.s. | n.s. | ↑ F, C | ↑ F, C, P, O, T |
| REM | ↑ F, C, P, O, T | ↑ F, C, P, O, T | n.s. | n.s. | ↑ F, C, P, O, T | ↑ F, C, P, O, T | n.s. | ↑ F, C, P, O, T | n.s. | n.s. | n.s. | n.s. |

*F – frontal, C – central, P – parietal, O – occipital, T – temporal electrodes, n.s. – non-significant, unmed. – unmedicated, med. – medicated, ↑ – increased compared to controls, REM – rapid eye movement sleep, N – non–rapid eye movement sleep.*
